# Few shot learning for phenotype-driven diagnosis of patients with rare genetic diseases

**DOI:** 10.1101/2022.12.07.22283238

**Authors:** Emily Alsentzer, Michelle M. Li, Shilpa N. Kobren, Ayush Noori, Undiagnosed Diseases Network, Isaac S. Kohane, Marinka Zitnik

## Abstract

There are over 7,000 rare diseases, some affecting 3,500 or fewer patients in the US. Due to clinicians’ limited experience with such diseases and the heterogeneity of clinical presentations, approximately 70% of individuals seeking a diagnosis remain undiagnosed. Deep learning has demonstrated success in aiding the diagnosis of common diseases. However, existing approaches require labeled datasets with thousands of diagnosed patients per disease. We present SHEPHERD, a few shot learning approach for multi-faceted rare disease diagnosis. SHEPHERD performs deep learning over a knowledge graph enriched with rare disease information and is trained primarily on simulated rare disease patients. We demonstrate SHEPHERD’s effectiveness across diverse diagnostic tasks, performing causal gene discovery, retrieving “patients-like-me”, and characterizing novel disease presentations, using real-world cohorts from the Undiagnosed Diseases Network (*N* = 465), MyGene2 (*N* = 146), and the Deciphering Developmental Disorders Study (*N* = 1, 431). SHEPHERD demonstrates the potential of knowledge-guided deep learning to accelerate rare disease diagnosis.

## Introduction

Rare diseases affect 300-400 million people worldwide, yet each disease has a very low prevalence, involving no more than 50 per 100,000 individuals [1–3]. Due to the low prevalence of rare diseases, most front-line clinicians lack firsthand experience, resulting in numerous specialty referrals and expensive clinical workups for patients across multiple years and institutions. Furthermore, patients with the same disease can present with variable symptoms, disease severity, and age of onset [4]. Such challenges make rare disease diagnosis extremely difficult; approximately 70% of individuals seeking a diagnosis remain undiagnosed, and the genes underlying up to 50% of Mendelian conditions are unknown [5,6]. These diagnostic delays can lead to redundant testing or unnecessary medical procedures, inappropriate or delayed disease management, and irreversible disease progression if the time window for intervention is missed.

Machine-assisted diagnosis offers the opportunity to shorten diagnostic delays for rare disease patients. Several strategies have been developed to automatically analyze patients’ genetic and phenotypic data to aid diagnosis. Genotype-based approaches focus on leveraging variant frequency and predicted pathogenicity to identify disease-causing variants [7–9]. Phenotype-based approaches prioritize genes by analyzing a patient’s facial image [10–15] or by comparing a patient’s set of phenotypic abnormalities to a knowledge base containing associations between phenotypes, genes, and diseases [16–20]. Other approaches combine genotype and phenotype-based approaches through Bayesian reasoning [21, 22] or by training machine learning models to combine multiple handcrafted features [23–28]. Automated diagnosis pipelines leveraging genotyping and phenotyping methods have improved diagnostic yields across a range of rare diseases [29, 30].

Advances in deep learning have considerably improved diagnostic accuracy in other clinical areas [31–40], achieving near-expert clinical accuracy for common diseases [41–43]. These methods offer several benefits: they can automatically learn valuable features from patient cohort data and integrate multimodal phenotypic and genomic data into a shared feature space. Despite the promise of deep learning approaches, however, there have been limited attempts to leverage deep learning methods for diagnosing rare genetic conditions due to the data scarcity problem. While deep learning approaches exist for image-based diagnosis [10, 11], there are limited approaches for phenotype-based diagnosis, and none automatically learn phenotypic and/or genotypic features over patient phenotype terms [23–28, 44]. Although foundation models, such as large language models, have been leveraged for clinical applications [45, 46], they are unable to achieve the diagnostic accuracy of traditional rare-disease decision support tools [47].

Training deep learning models requires high-quality labeled data with thousands of diagnosed patients per disease. Yet, rare disease datasets are three orders of magnitude smaller. The low prevalence of rare diseases makes it difficult to obtain datasets of sufficient size for deep learning, even with manual expert curation. Training deep learning models on datasets with smaller sample sizes can impact their generalizability. Due to each rare disease’s heterogeneity and low prevalence, models are unlikely to have seen patients with the same (or similar) genetic disorders during training. Deep learning approaches for rare disease diagnosis must be able to extrapolate beyond the training distribution to novel genetic conditions and atypical disease presentations. Deep learning approaches that leverage medical knowledge are needed to overcome the limitations of traditional supervised deep learning methods in this limited data setting.

Here, we introduce SHEPHERD, a deep learning approach for multi-faceted diagnosis of patients with rare genetic conditions. SHEPHERD inputs patient phenotype terms and an optional list of candidate genes, and operates at multiple points throughout the rare disease diagnosis process to perform causal gene discovery, retrieve “patients-like-me” with similar genetic and phenotypic features, and provide interpretable names for novel disease presentations. To overcome the limitations of supervised learning, SHEPHERD performs label-efficient training by (1) training primarily on simulated rare disease patients and (2) incorporating medical knowledge of known phenotype, gene, and disease associations via knowledge-guided deep learning. The simulated patients are created using an adaptive simulation approach that generates realistic rare disease patients with varying numbers of phenotype terms and candidate genes [48]. Knowledge-guided learning is achieved by training a graph neural network to represent a patient (specifically, the patient’s presenting phenotypic features) about other phenotypes, genes, and diseases. When a new patient arrives, SHEPHERD produces a mathematical representation (*i*.*e*., embedding) of the patient in the latent space such that the patient is embedded near the patient’s causal gene(s), disease, and other patients with the same causal gene(s) or disease, and far from irrelevant genes and diseases and other patients with different diseases. Using SHEPHERD’s embedding space optimized for rare disease diagnosis, SHEPHERD can nominate genes and diseases for every patient, even when no other patients are known to be diagnosed with the same disease. Unlike existing methods, which rely on handcrafted features, SHEPHERD learns patient representations informed by medical knowledge directly from patient phenotype terms to enable rare disease diagnosis with few (or zero) labeled examples.

We evaluate SHEPHERD on an external cohort of patients in the Undiagnosed Diseases Network (UDN) [49], a nationwide initiative with 12 clinical sites in the US tasked with diagnosing patients with rare, difficult-to-diagnose genetic conditions. In addition to the multi-site UDN cohort, our external evaluation includes a nationwide MyGene2 patient cohort. SHEPHERD performs granular, phenotype-based causal gene discovery by ranking candidate genes from bioinformatics pipelines. We find that SHEPHERD ranks the correct gene first in 40% of patients spanning 16 disease areas, improving diagnostic efficiency by at least twofold compared to a non-guided baseline. In addition, SHEPHERD nominates the correct diagnosis for patients with atypical presentations or novel diseases, ranking the correct gene among the top five predictions for 77.8% of those hard-to-diagnose patients. SHEPHERD excels in diagnosing patients with novel genetic conditions, ranking up to 86% of patients the same as or better than domain-specific approaches. By testing SHEPHERD on each disease area, clinical site, and year of diagnosis, we find that SHEPHERD has sustained performance over time and across diseases and clinical sites in the UDN. Further, SHEPHERD generates patient representations that capture patient similarity (Adjusted Mutual Information = 0.304) and enable retrieval of “patients-like-me” with similar genetic conditions. Finally, SHEPHERD can provide interpretable characterizations of novel disease presentations. By describing novel diseases based on their similarity to known diseases, SHEPHERD can point clinical researchers towards the most closely related diseases to investigate the novel disease in depth. For each diagnostic task, we illustrate SHEPHERD’s capabilities on patients in the Undiagnosed Diseases Network and provide an interactive tool to explore SHEPHERD’s predictions at https://huggingface.co/spaces/emilyalsentzer/SHEPHERD.

## Results

### Overview of the Undiagnosed Diseases Network patient cohort

We assemble a cohort of 465 patients in the Undiagnosed Diseases Network (UDN) with molecular diagnoses. Most patients are diagnosed with a single causal gene that explains their symptoms; 14 patients (3%) have two causal genes, and two patients (0.4%) have three causal genes. Most patients in the UDN receive an extensive clinical workup and whole genome or exome sequencing (Figure 1a). Sequencing data is analyzed with the involvement of clinicians and genetic counselors to identify candidate genes that harbor variants likely to explain the patient’s symptoms. Once one to five strong candidates are identified, causality is assessed by searching for genotype- and phenotype-matched individuals in human and animal databases or by introducing candidates into model organisms to determine *in vivo* impact [50].

**Figure 1:**
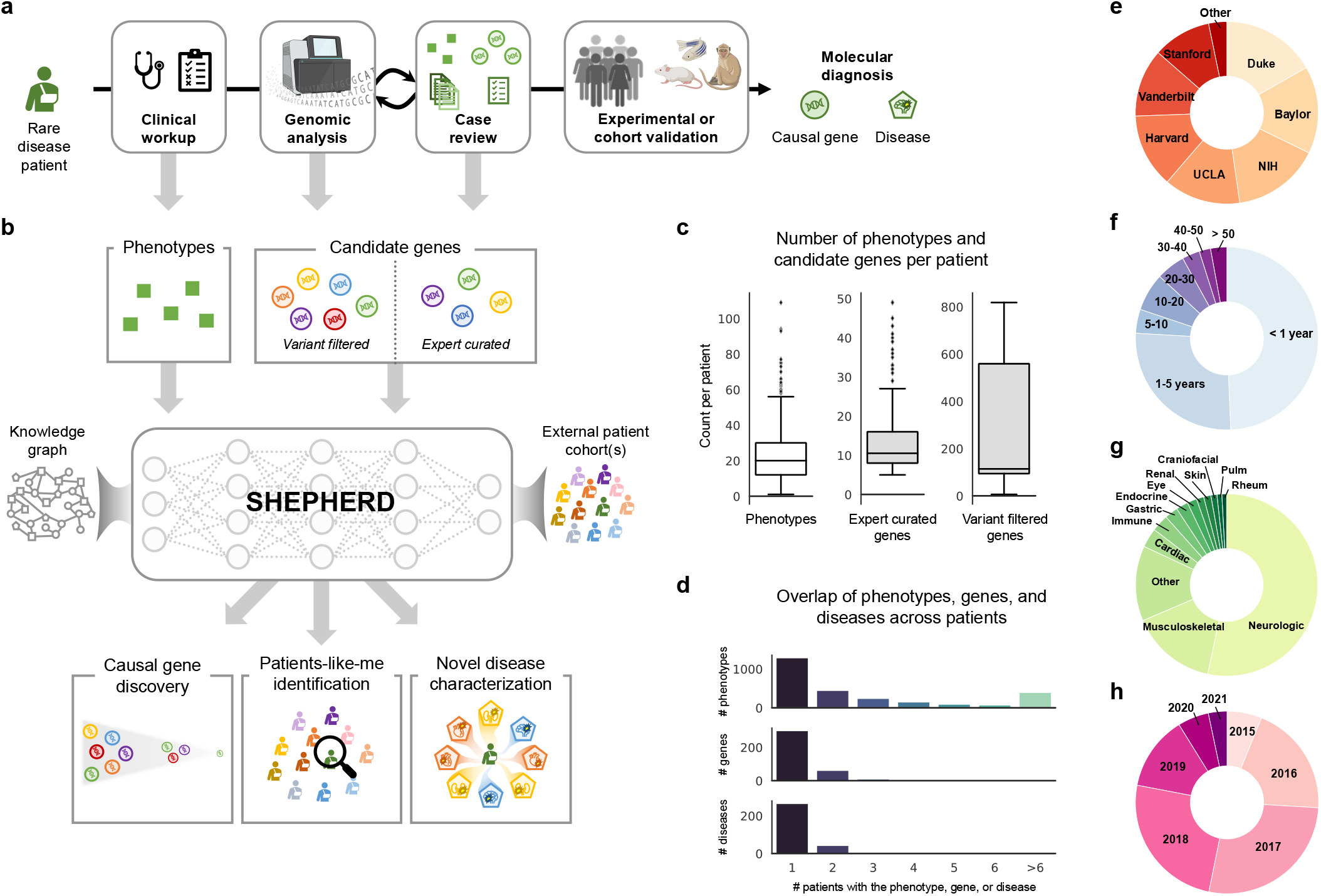
Overview of SHEPHERD in the rare disease diagnosis pipeline. **(a)** After years of failed diagnostic attempts, once a patient is accepted to the UDN, they receive a thorough clinical workup and genetic sequencing, and their case is analyzed in an iterative process to identify the candidate genes likely to explain the patient’s symptoms. SHEPHERD can be used throughout the diagnostic process: after the clinical workup to find similar patients, after the sequencing analysis to identify strong candidate genes, and after the case review to further prioritize candidate genes, characterize the patient’s disease, and/or validate candidate genes by finding phenotype- and genotype-matched patients. **(b)** SHEPHERD takes in as input the patient’s set of phenotype terms and leverages an external rare disease knowledge graph to perform multi-faceted rare disease diagnosis. SHEPHERD can optionally consider a list of candidate genes (either variant-filtered or expert-curated) or external patient cohort(s), depending on the prediction task of interest (e.g., causal gene discovery, patients-like-me identification). For simplicity, the knowledge graph is depicted using three shapes: circles as genes, squares as phenotypes, and pentagons as diseases; refer to Methods for all node types. **(c)** Number of HPO phenotype terms and candidate genes in each of the two candidate gene lists across patients in our UDN cohort. **(d)** Overlap of phenotype terms, genes, and diseases across patients. Most phenotype terms, genes, and diseases are found in only a single UDN patient. **(e-h)** Number of patients in each **(e)** UDN clinical site, **(f)** age category, **(g)** primary presenting symptom, and **(h)** evaluation year. Figure adapted from images created in BioRender [91].

Through this diagnostic process, patients are annotated with a set of Human Phenotype Ontology (HPO) phenotype terms describing their clinical features and a set of candidate genes that may explain the patient’s syndrome. Clinical experts additionally annotate diagnosed patients with an Online Mendelian Inheritance in Man (OMIM) identifier describing their disease (if available). Each patient is characterized by 23.9 HPO terms on average (SD = 16.1; Figure 1c). The candidate genes are patient-specific and include genes in which the patient has a mutation. For each patient, the diagnostic process creates two sets of candidate gene lists. The lists contain genes considered at two different phases in the UDN diagnosis pipeline (Figure 1a): VARIANT-FILTERED, a list produced by performing initial variant-based filtering of candidate genes, and EXPERT-CURATED, a list including genes marked by clinical experts as strong candidates for the patient (Methods 3). The VARIANT-FILTERED gene lists are produced using Exomiser [24, 51], a variant-based tool used in parallel to existing pipelines at three UDN sites [50]. The two candidate gene lists contain 244.3 and 13.3 genes on average, respectively (SD = 244.0 and SD = 8.0; Figure 1c). Each gene list is input to SHEPHERD to predict the causal gene (*i*.*e*., the gene harboring variants that cause the patient’s disease) from both a long list of candidate genes derived from automated filtering (*i*.*e*., VARIANT-FILTERED) and a short list of the strongest candidate genes that are more challenging to prioritize (*i*.*e*., EXPERT-CURATED).

UDN patients have heterogeneous disease presentations: 378 unique genes and 299 unique diseases are represented in the cohort, and 48% of phenotype terms, 79% of genes, and 83% of diseases are represented in only a single patient (Figure 1d). This reinforces the need for machine learning models that can learn from sparsely labeled datasets. 11.4% patients have a diagnosis in common with at least one other patient. Patients with the same disease have only 67% of phenotype terms in common on average (SD = 43%), and the closest shared ancestor (*i*.*e*., lowest common ancestor) in the Human Phenotype Ontology between their phenotype terms is 2.67 hops away on average (SD = 0.81). In addition, 7% of patients have novel genetic diseases, and only 28% of each patient’s phenotypic features have any known association with the causal gene on average (SD = 21%). The assembled cohort of UDN patients has been evaluated at 12 clinical sites across the United States (Figure 1e). While 75.9% of patients are under five years old, patients can present to the UDN with suspected genetic diseases in their 40s or 50s (Figure 1f). The cohort is predominantly White (80.6%) and non-Hispanic (70.8%); smaller proportions of patients identify as Asian (9.2%), Black or African American (4.5%), or other racial and ethnic backgrounds (5.6%; Supplementary Figure 1a,b). The sex distribution is relatively balanced, with 47.7% male and 52.0% female patients (Supplementary Figure 1c). Most patients present with neurological symptoms but can exhibit cardiac, musculoskeletal, rheumatic, and many other symptoms (Figure 1g). Due to the lag between starting the process at the UDN and receiving the diagnosis, most patients included in the analysis were evaluated by UDN clinicians in 2016-2018 (Figure 1h). The phenotypic heterogeneity and presence of novel and atypical diseases pose a challenge for diagnosis, requiring diagnostic technology that can accommodate previously unseen phenotypes, genes, and diseases and leverage knowledge beyond direct gene, phenotype, and disease associations (Supplementary Figure 2). The UDN patients represent a diverse, independent cohort used exclusively for model evaluation. Importantly, these patients are not used to train SHEPHERD.

### Overview of SHEPHERD algorithm

SHEPHERD takes a set of patient’s phenotype terms and candidate disease(s) or candidate gene(s) harboring causal variants as input, and performs multi-faceted diagnosis of the patient to identify causal genes, retrieve “patients-like-me” with the same causal gene or disease, and provide interpretable characterizations of novel disease presentations (Figure 1b). SHEPHERD can integrate into the rare disease diagnostic process workflow at multiple points: (1) to find similar patients after the patient’s clinical workup, (2) to identify strong candidate causal genes after the initial sequencing analysis or in conjunction with the clinical case review, and (3) to characterize the patient’s disease and find similar patients for experimental or cohort validation after candidate causal genes are identified (Figure 1a-b).

SHEPHERD is a few shot geometric deep learning approach for rare disease diagnosis. Fewshot learning, which can make predictions when very few (if any) labeled data points are available, is central to rare disease diagnosis because of the low prevalence of each disease. Key to SHEPHERD’s ability to provide diagnostic prediction when zero or at most a few labeled (diagnosed) patients per disease are available is to use a biomedical knowledge graph containing gene, phenotype, and disease relationships. SHEPHERD represents each patient as a set of phenotype terms from the knowledge graph, which we refer to as a phenotype subgraph to emphasize that these terms are embedded within the graph’s structure (Methods 1). It leverages a graph neural network to jointly embed each patient’s phenotype subgraph and candidate genes or diseases into a latent representation space such that the generated embeddings are informed by the structure of the knowledge graph, and patients are embedded nearby their causal gene(s), disease(s), and other similar patients (Figure 2a-b). Further, SHEPHERD uses an attention mechanism to aggregate each patient’s phenotype terms to generate a patient embedding. While not intended as a clinical interpretability tool, the attention weights can be inspected post hoc to probe how the model prioritizes different phenotypic features during training and inference.

**Figure 2:**
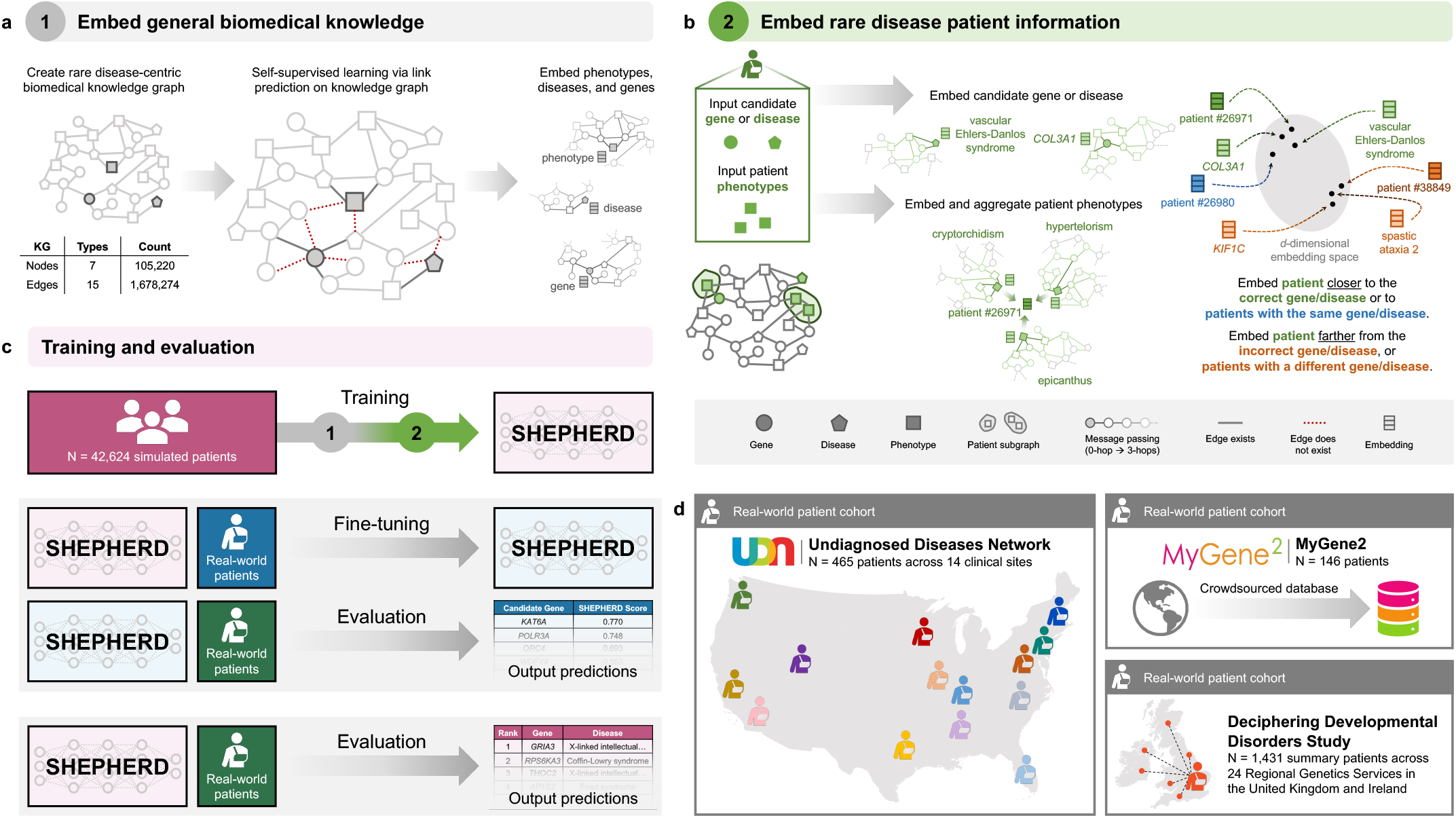
SHEPHERD architecture, training, and generalizability. **(a-b)** SHEPHERD is trained in a two-step process. **(a)** First, the model is pretrained to embed the biomedical knowledge in the knowledge graph. **(b)** Then, the pretrained model is applied to the task of rare disease diagnosis. Patient information is overlaid on the knowledge graph, and SHEPHERD generates an embedding for the patient phenotype terms and each candidate gene, disease, or patient. The model is trained via a loss function that encourages patient embeddings to be close to the embeddings of their causal gene or disease or other patients with the same causal gene or disease. **(c)** SHEPHERD is trained on a large cohort of simulated patients (pink). It can be further trained on real-world patients (blue) and then evaluated on an independent cohort of real-world patients (green). Alternatively, SHEPHERD can directly be evaluated on real-world patients (green) without any additional training. **(d)** We leverage real patient data derived from three distinct cohorts: the Undiagnosed Diseases Network (UDN; *N* = 465), MyGene2 (*N* = 146), and Deciphering Developmental Disorders Study (DDD; *N* = 1, 431). For simplicity, the KG is depicted using three shapes: circles as genes, squares as phenotypes, and pentagons as diseases; refer to Methods for all node types.

SHEPHERD is trained in a two-step process to learn embeddings of biomedical concepts and patients with rare genetic diseases. First, SHEPHERD is pretrained via self-supervised learning to embed genes, phenotypes, and diseases by predicting the relationships (structure) of the biomedical knowledge graph (Figure 2a; Methods 7). This step produces compact embeddings that can be adapted for a range of analyses and are generalizable by accounting for complementarity between diseases. Then, using the pretrained model as initialization, SHEPHERD is trained for multi-faceted diagnosis of patients with rare diseases via a novel objective function (Figure 2b; Methods 7). We train SHEPHERD in a disease-stratified manner (*i*.*e*., in which patients with the same disease are assigned either to the training or validation set, but not both) to enable SHEPHERD to generalize to diseases unseen during training.

Due to the scarcity of data for patients with rare diseases, we leverage simulated but realistic rare disease patients for training SHEPHERD (Figure 2c). We train SHEPHERD on a cohort of more than 40,000 synthetic rare disease patients representing over 2,000 rare diseases in Orphanet (Methods 6). There are 20 synthetic patients generated for each rare disease. The simulated patients were generated using an approach designed to generate realistic rare disease patients grounded in medical knowledge, and they have been shown to phenotypically and genetically resemble real-world rare disease patients [48]. The synthetic cohort is essential for training SHEPHERD, as it is considerably larger, more diverse, and more representative of phenotype and genotype heterogeneity than any real-world dataset of rare disease patients (Figure 2c) [48]. This dataset, together with knowledge-guided learning on the rare disease knowledge graph, enables deep learning for rare disease diagnosis. A notable byproduct of training the model on synthetic data is that SHEPHERD’s model can be publicly released without the risk of exposing patient information [52]. After training, SHEPHERD can be further trained on real-world patient cohorts or leveraged directly for rare disease diagnosis.

We leverage real patient data from three cohorts in this study (Figure 2d): (1) the UDN patient cohort (Methods 3); (2) a cohort of 146 patients from MyGene2, an online portal through which families with rare genetic conditions can share their health information to connect with clinicians and other patients [53] (Methods 4); (3) a cohort of 1,431 patients derived from the Deciphering Developmental Disorders Study, an initiative from the United Kingdom and Ireland designed to diagnose patients with undiagnosed developmental disorders [54] (Methods 5). Results are described in the following sections.

### SHEPHERD can perform causal gene discovery

A critical step in rare disease diagnosis is identifying the gene(s) that are strong candidates for causing the patient’s syndrome (Figure 1a). Given a patient’s set of phenotype terms and a list of genes in which the patient has a mutation, SHEPHERD predicts genes that harbor variants most likely to explain the patient’s presenting symptoms. SHEPHERD produces a score for each candidate gene in the patient that fuses two complementary aspects of information: an embedding-based metric that captures the global network topology and a network-based metric computed using knowledge graph distance that captures local network information (Methods 11). We use SHEPHERD to prioritize genes found in both the EXPERT-CURATED and VARIANT-FILTERED candidate gene lists (Methods 3). In both instances, SHEPHERD performs granular prioritization by refining lists of patients’ candidate genes output by bioinformatics pipelines. For this analysis, we leverage patients from three cohorts: the simulated, MyGene2, and DDD cohorts are used for training, and the UDN cohort is used for validation.

We report SHEPHERD’s performance in causal gene discovery as the average recall at *k*, defined as the number of causal genes correctly predicted in the top *k* ranked genes on average for all patients in the cohort. On the EXPERT-CURATED gene lists, SHEPHERD ranks the patient’s causal gene first in 40% of UDN patients, achieving a recall of 0.69 when *k* = 3 and 0.85 when *k* = 5 on average (Figure 3a). On the much longer VARIANT-FILTERED gene lists, SHEPHERD achieves an average recall of 0.21, 0.38, and 0.48 for *k* = 1, 5, and 10, respectively (Figure 3d).

**Figure 3:**
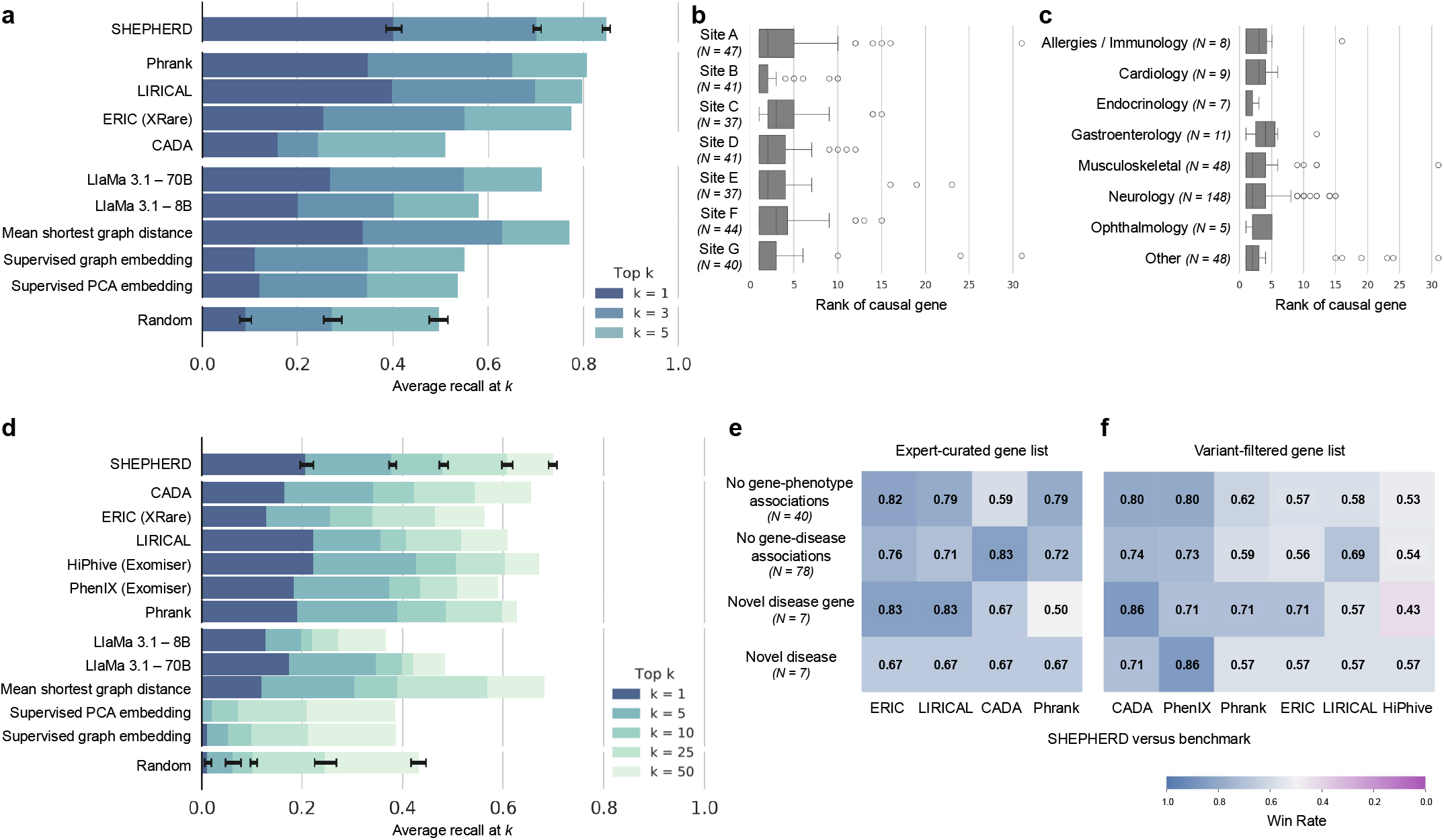
SHEPHERD performs generalizable causal gene discovery. **(a)** Performance of SHEPHERD, four domain-specific approaches, five language model, traditional machine learning, and network science baselines, and a random baseline. The performance metric is average recall at *k* for *k* = 1, 3, and 5. Error bars denote standard deviation over models trained with 5 random seeds. (b-c) Performance of SHEPHERD in ranking causal genes stratified by **(b)** clinical sites and **(c)** primary presenting symptoms. Each boxplot shows the median and interquartile range of the rank of the causal gene. Whiskers extend to ±1.5 × IQR. **(d)** Performance of SHEPHERD, six domain-specific approaches, five large language model, traditional machine learning, network science baselines, and a random baseline. The performance metric is average recall at *k* for *k* = 1, 5, 10, 25, and 50. Error bars denote standard deviation over models trained with 5 random seeds. **(e-f)** Performance of SHEPHERD against domain-specific algorithms in four extremely hard-to-diagnose scenarios on **(e)** EXPERT-CURATED and **(f)** VARIANT-FILTERED gene lists. Shown is the win rate, the proportion of patients where SHEPHERD performs the same as or better than the benchmark algorithms.

We find no significant difference in performance across UDN sites throughout the US, patients with varying presenting symptoms, and the year of evaluation by UDN clinicians (Figure 3b-c, Supplementary Figure 3a, Supplementary Figure 4a-c) on both the EXPERT-CURATED and VARIANT-FILTERED gene lists. These results indicate that SHEPHERD can generalize across clinical sites and diseases over time. Furthermore, we find that SHEPHERD’s performance does not correlate with the number of annotated phenotype terms for each patient (Spearman’s *ρ* = 0.02 and *ρ* = *−*0.11 for EXPERT-CURATED and VARIANT-FILTERED lists respectively; Supplementary Figure 3c and Supplementary Figure 4e). Finally, we evaluate SHEPHERD’s performance as a function of the prevalence of the rare disease. We leverage the number of submissions to ClinVar as a proxy for prevalence. We find that SHEPHERD’s performance does not strongly correlate with the prevalence of the genetic condition (Spearman’s *ρ* = *−*0.17 and *ρ* = *−*0.16 for EXPERT-CURATED and VARIANT-FILTERED lists respectively; Supplementary Figure 3d and Supplementary Figure 4f). SHEPHERD’s ability to generalize represents an important capability because rare disease patients are heterogeneous, and developing separate predictive models that perform well for each patient subgroup is not feasible due to the low prevalence of the disorders.

**Figure 4:**
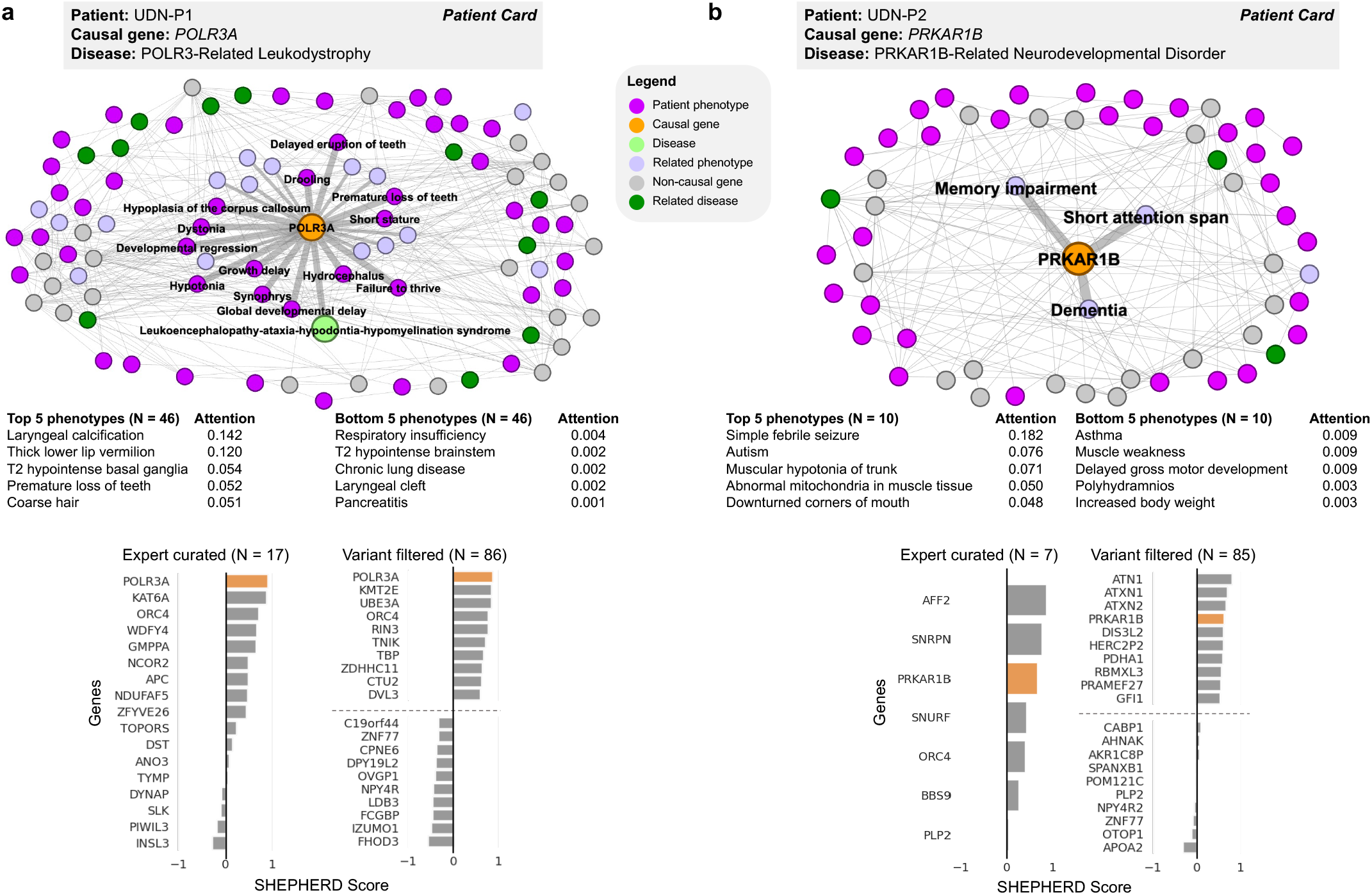
Causal gene discovery case studies for patients with novel genetic conditions. SHEPHERD identifies the causal gene even in atypical or novel disease presentations. Each patient case study, shown in **(a)** and **(b)**, includes the subset of the knowledge graph containing all nodes in the shortest path between the patient’s phenotype terms, causal gene, and disease; a table of the patient’s phenotype terms and attention weights learned by SHEPHERD; and bar plots of scores SHEPHERD assigned to each candidate gene in the EXPERT-CURATED and VARIANT-FILTERED lists. The top and bottom 5 ranked genes in the VARIANT-FILTERED list are shown. The causal gene is highlighted in orange. The direct phenotypic neighbors of the causal gene are emphasized. In patient UDN-P1’s network, the patient’s causal gene is directly connected to the disease in the knowledge graph. In patient UDN-P2’s network, there is no disease node because the patient has a novel uncharacterized syndrome. All panels, except those labeled as a “Patient Card” (colored box with the information provided by the UDN), depict SHEPHERD’s predictions or analyses performed on outputs of SHEPHERD.

We evaluate SHEPHERD against 12 baseline approaches (Methods 21). We select a network science algorithm and two supervised machine learning approaches as benchmarks to quantify the advantages of SHEPHERD’s graph neural network approach. We also identify six domain-specific algorithms developed for causal gene discovery that leverage information theory (Phrank [16], PhenIX [24], and ERIC [25]), likelihood ratios (LIRICAL [21]), shallow graph embeddings (CADA [19]), and information-theoretic and random walk methods (HiPhive [24]). We further evaluate SHEPHERD against two large language models (LlaMa 3.1 8B and 70B [55]; Supplementary Figure 8). SHEPHERD performs comparably or significantly better than all benchmarking approaches on the EXPERT-CURATED and VARIANT-FILTERED gene lists (Figure 3a,d; Supplementary Figure 8). SHEPHERD outperforms the strongest domain-specific algorithms, LIRICAL and HiPhive, in prioritizing causal genes overall on both EXPERT-CURATED (*p*-value = 4.27 *×* 10^*−*2^ for LIRICAL) and VARIANT-FILTERED (*p*-value = 2.05 *×* 10^*−*4^ for LIRICAL and *p*-value = 2.70 *×* 10^*−*5^ for HiPhive) gene lists (Wilcoxon signed rank sum test). SHEPHERD significantly outperforms the other domain-specific approaches in retrieving the causal gene first by up to 24.4% (*p*-value = 4.92 *×* 10^*−*16^) and 7.7% (*p*-value = 1.55 *×* 10^*−*3^) of patients on the EXPERT-CURATED and VARIANT-FILTERED gene lists, respectively (McNemar’s test). Furthermore, SHEPHERD significantly outperforms large language models in retrieving the causal gene first by up to 20.1% (*p*-value = 1.42 *×* 10^*−*9^) and 7.9% (*p*-value = 6.85 *×* 10^*−*3^) of patients on the EXPERT-CURATED and VARIANT-FILTERED gene lists, respectively, and the other machine learning approaches by up to 29.0% (*p*-value 1.73 *×* 10^*−*17^) and 20.4% (*p*-value 1.44 *×* 10^*−*15^) of patients, respectively (McNemar’s test). For these statistical tests, we apply Benjamin-Hochberg procedure for multiple testing correction.

SHEPHERD’s strong performance demonstrates that SHEPHERD can complement existing variant-based approaches for gene prioritization while leveraging the extensive knowledge sources of gene-phenotype associations. Using SHEPHERD, rare disease experts would need to evaluate 1,026 genes from the EXPERT-CURATED lists or 18,005 genes from the VARIANT-FILTERED lists to arrive at the causal gene for all 465 UDN patients. In contrast, with non-guided ranking, experts would need to evaluate a total of 2,231 EXPERT-CURATED genes or 27,727 VARIANT-FILTERED genes, suggesting that SHEPHERD has the potential to improve diagnostic efficiency by 2.2-times and 1.5-times, respectively. Compared to the best domain-specific approaches, LIRICAL and HiPhive, SHEPHERD reduces the number of genes that experts need to consider by 97 (8.6%) and 5,495 (23.3%) on the EXPERT-CURATED and VARIANT-FILTERED gene lists, respectively (LIRICAL), and by 1,878 (9.4%) on the VARIANT-FILTERED gene list (HiPhive).

### SHEPHERD can diagnose patients with atypical and novel genetic diseases

Patients in the UDN have atypical or novel disease presentations, which makes them challenging to diagnose because there are no direct associations between patients’ genes, symptoms, and the correct diagnosis. Consequently, the lack of direct linkage between patients’ phenotypic features and the correct diagnosis (causal genes) means that a lookup against medical knowledge bases is ineffective for diagnosis. We find that SHEPHERD can identify the causal gene even when the patient’s presenting phenotypic abnormalities are multiple hops away from the gene causing the disease in the knowledge graph. For 77.8% of patients whose phenotype terms are far away from their causal genes in the knowledge graph (i.e., more than two hops away), SHEPHERD identifies the correct causal gene among its top five predictions from the EXPERT-CURATED gene list. No strong correlation exists between SHEPHERD’s performance and the distance between the patient’s phenotype terms and causal gene (Supplementary Figure 3b, Supplementary Figure 4d; *R*^2^ = 0.102, Spearman’s *ρ* = 0.37 and *R*^2^ = 0.0004, Spearman’s *ρ* = 0.12 for the EXPERT-CURATED and VARIANT-FILTERED gene lists, respectively).

We evaluate SHEPHERD against the domain-specific models in four hard-to-diagnose scenarios (Figure 3e). We identify patients from the UDN whose causal genes lack known associations with phenotype terms or diseases in the literature (based on our rare disease knowledge graph) and who have been identified by UDN experts as having novel disease genes or novel diseases. SHEPHERD achieves win rates (*i*.*e*., ranks the causal gene the same or higher) of up to 82% and 83% for patients whose causal genes have no known phenotype or disease associations, respectively, on the EXPERT-CURATED gene lists. On the VARIANT-FILTERED gene lists, the win rates are up to 80% and 74%, respectively. SHEPHERD achieves win rates of up to 67% and 83% for patients with a novel disease or novel disease gene, respectively, according to UDN experts on the EXPERT-CURATED gene lists, and up to 86% on the VARIANT-FILTERED gene lists. The only subset of patients for which a baseline performs slightly better than SHEPHERD consists of patients with novel disease genes, according to human experts in the UDN. In all other scenarios, SHEPHERD outperforms all baseline approaches, demonstrating SHEPHERD’s ability to diagnose patients with atypical and novel genetic diseases.

We further demonstrate the use of SHEPHERD for patients diagnosed with an atypical presentation of a known disease or a novel syndrome through two case studies on patients from the UDN. Patient UDN-P1 (Figure 4a; SHEPHERD Tool Tab 1, Patient UDN-P1) received a diagnosis for POLR3-related leukodystrophy three years after acceptance into the UDN. While the involvement of gene *POLR3A* with leukodystrophy (MIM:607694) is known, the patient’s case was challenging due to her atypical clinical presentation. Several of her presenting clinical features, including lack of tear production, premature adrenarche, laryngeal cleft, hearing loss, and high blood pressure, are not typical of leukodystrophy. Further, only 28.3% (13 out of 46) of the patient’s phenotype terms are directly linked to *POLR3A* in the knowledge graph, and the patient phenotype terms are 1.98 hops away from the causal gene in the knowledge graph on average. The *POLR3A* gene is associated with five other diseases, and 93.7% (192 out of 205) of phenotype terms directly linked to *POLR3A* are not found in the patient, further complicating the diagnosis. Despite this atypical disease presentation, SHEPHERD identifies the patient’s causal gene in the top 1 out of 17 and 86 candidate genes in the EXPERT-CURATED and VARIANT-FILTERED gene lists, respectively. Strikingly, SHEPHERD can disambiguate diseases by optimally up- and down-weighting phenotypic features using an attention mechanism, and correctly down-weights phenotypic features that are atypical of leukodystrophy.

SHEPHERD can also identify strong candidate genes for patients with novel uncharacterized syndromes. Patient UDN-P2 (Figure 4b; SHEPHERD Tool Tab 1, Patient UDN-P2) was accepted into the UDN with congenital hypotonia and developmental delay. While no diagnosis was identified in the primary genomic and clinical evaluation, the patient was diagnosed three years later with a novel *PRKAR1B*-related neurodevelopmental disorder. The *PRKAR1B* gene is not associated with known diseases. None of the 21 phenotype terms directly linked to *PRKAR1B* are found in the patient, and the average shortest path length from the patient’s phenotype terms to the causal gene is 2.4. Nevertheless, SHEPHERD identifies the suspected causal gene among the top 3 in the EXPERT-CURATED candidate list and the top 4 in the VARIANT-FILTERED candidate list, illustrating how SHEPHERD can assist in recognizing novel genetic diseases.

### SHEPHERD learns meaningful patient representations that capture patient similarity

Another critical consideration for rare disease diagnosis is finding patients that share the same disease or causal gene, commonly referred to as “patients-like-me” [56] (Figure 1a). Starting from a set of patient phenotype terms, SHEPHERD flags other patients in the cohort with similar genetic diseases suitable for follow-up diagnostic analysis. Concretely, SHEPHERD finds similar patients through a deep embedding scorer optimized to represent patients with the same causal genes or disease as nearby points in the embedding space. For this analysis, we leverage patients from three cohorts: the simulated cohort is used for training, and the UDN and MyGene2 cohorts are used for validation.

SHEPHERD represents each patient as a point in the embedding space colored by the disease category of their diagnosed disease (Figure 5). The categories correspond to the 33 disease categories outlined in Orphanet (Methods 2). Robust clustering of patients by disease area (AMI = 0.304; *p*-value *<* 0.01) shows that SHEPHERD generates an embedding space that meaningfully captures patient relationships that can directly answer “patients-like-me” queries. Remarkably, even though SHEPHERD is trained on simulated patients, it generalizes to real-world UDN and MyGene2 cohorts, revealing disease-enriched regions in the embedding space where real-world patients are positioned nearby simulated patients with the same disease area (Supplementary Figure 5).

**Figure 5:**
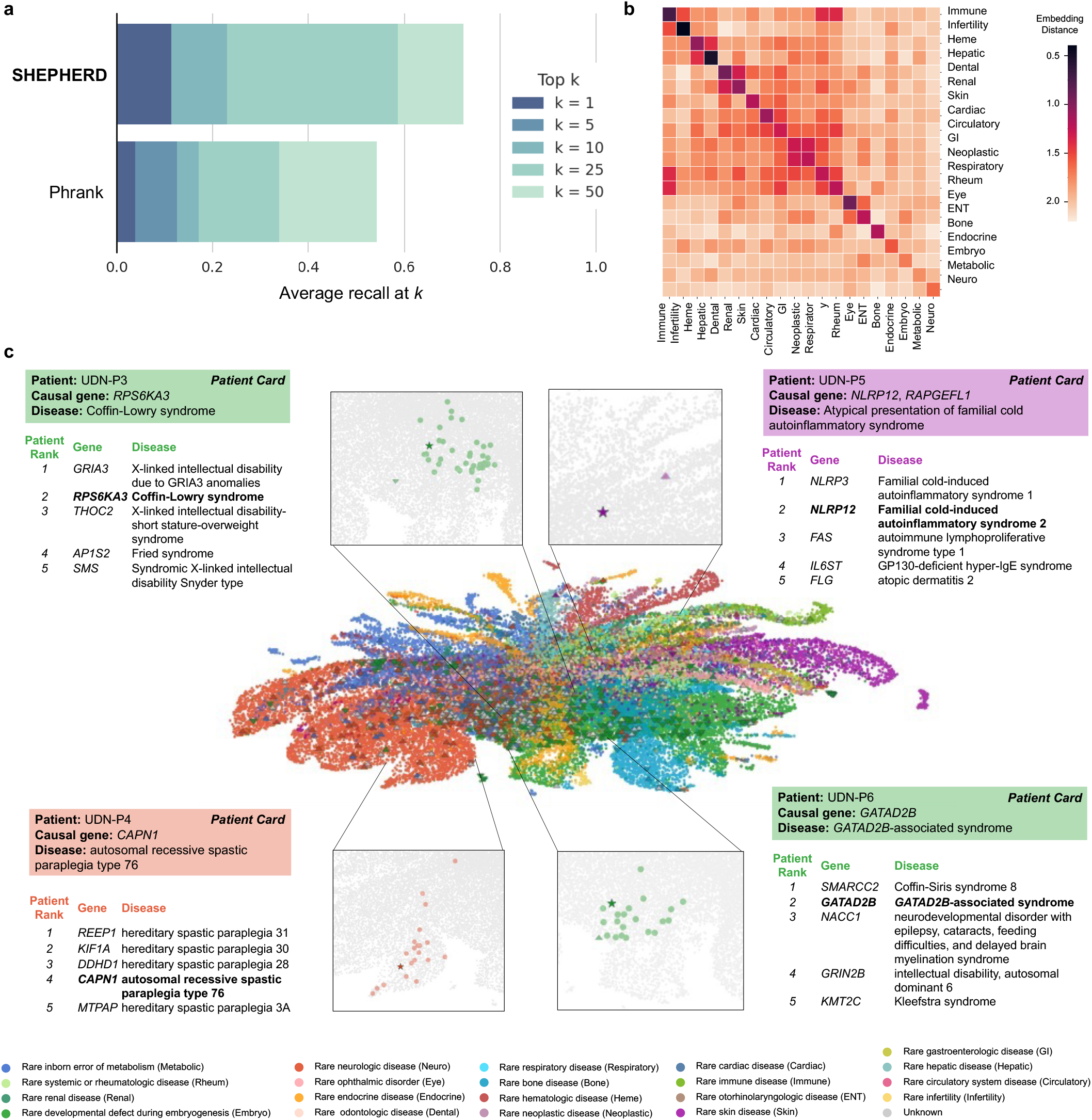
SHEPHERD identifies patients-like-me from simulated, UDN, and MyGene2 cohorts. **(a)** Performance of SHEPHERD in retrieving MyGene2 patients with the same causal gene as a UDN patient (*n*=75 UDN patients with at least one matching patient in the MyGene2 cohort). SHEPHERD is benchmarked against Phrank, a domain-specific algorithm. The performance metric is average recall at *k* for *k* = 1, 5, 10, 25 and 50. **(b)** Heatmap of the average distance between the phenotype embeddings of pairs of patients across disease ca2t5egories. Darker colors indicate smaller distances and lighter colors indicate larger distances between patients of each pair of disease categories. **(c)** Two-dimensional UMAP plot of SHEPHERD’s embedding space of all simulated (circle), UDN (up-facing triangle), and MyGene2 (down-facing triangle) patients colored by their Orphanet disease category. Each of the four case studies consists of a zoomed-in UMAP displaying the query patient (star) and all patients with the same causal gene as the query (colored circles) and a table containing information regarding the top five most similar patients retrieved by SHEPHERD. Patients are bolded in the table if they share the same causal gene. All panels, except those labeled as a “Patient Card” (colored box with the information provided by the UDN), depict SHEPHERD’s predictions or analyses performed on outputs of SHEPHERD.

To further evaluate patient embeddings, we compare embedding distances between patients diagnosed with either the same or different disease (*i*.*e*., comparing diagonal vs. off-diagonal entries, Figure 5b). We find that distances between patients of the same category are significantly smaller than between patients of different categories (*p*-value *<* 0.001 across all disease categories; Mann-Whitney test with Benjamin-Hochberg procedure), which indicates that SHEPHERD captures the similarity between patients with similar disease presentations. We also observe several distinct clusters of disease categories in the embedding space (Figure 5b; Supplementary Figure 7). For example, patients with neoplastic diseases and gastroenterologic diseases cluster together. Similarly, patients with hematologic and hepatic diseases and patients with odontologic and renal diseases cluster together in the embedding space. These clusters represent real co-occurrences of symptoms in disease presentations. For instance, patients with odontologic diseases, atypical dentin dysplasia, and orofaciodigital syndrome I, have both orofacial and renal disease presentations. Atypical dentin dysplasia is caused by a mutation in *SMOC2*, a matricellular protein involved in both craniofacial development and kidney fibrosis [57,58]. Orofaciodigital syndrome I is caused by a mutation in *OFD1*, which is involved in organogenesis and plays a vital role in the normal growth of orofacial and kidney tissues [59, 60]. These relationships reflect that diseases often involve multiple organ systems and indicate that the embedding space can capture the relationship between patients with similar symptoms even when their diagnoses differ.

### SHEPHERD can identify “patients-like-me” with similar genetic diseases

We next examine SHEPHERD’s ability to identify “patients-like-me” from a large cohort of rare disease patients. We either rank all simulated, UDN, and MyGene2 patients (UDN-P3 and UDN-P4 cases) or all UDN and MyGene2 patients (UDN-P5 and UDN-P6 cases; Figure 5c; SHEPHERD Tool Tab 2) to identify patients most similar to the query UDN patient. We locate each query patient and all similar patients with the same causal gene in SHEPHERD’s embedding space, and find that patients with the same causal gene are embedded nearby. In all four patient cases, SHEPHERD retrieves patients with the same causal gene and disease as the query patient among the top 5 predictions. Patients ranked above the patient with the same causal gene have very similar disease presentations to the query patient. For UDN-P4 and UDN-P5, the patients have a variant of the same disease caused by a different gene (Figure 5c). For UDN-P6, patients with Coffin-Siris syndrome 8 (ranked first) and *GATAD2B*-associated syndrome (ranked second) both exhibit impaired intellectual development, hypotonia, feeding difficulties, and hypertelorism, among other phenotypic abnormalities. For UDN-P3, patients with X-linked intellectual disability due to *GRIA3* (ranked first) and Coffin-Lowry syndrome (ranked second) share impaired intellectual development, seizures, scoliosis, and other phenotypic abnormalities.

The most similar patients identified by SHEPHERD do not necessarily have the most phenotype terms in common with the query patient. This reflects SHEPHERD’s ability to capture phenotypic similarity rather than just calculating a direct overlap in phenotype terms, typical of some information-theoretic approaches used in practice. In particular, patients that share the same causal gene have two to four phenotype terms in common. Only 10.0%, 9.0%, 26.6%, and 7.7% of the phenotype terms found in query patients UDN-P3, UDN-P4, UDN-P5, and UDN-P6 are also found in the most similar genotype-matched individual respectively. In contrast, patients with the most phenotype terms in common with the query are ranked at positions 366, 463, 41, and 16, respectively. For example, one patient shares 10 phenotype terms with UDN-P6, which is 38.5% of UDN-P6’s phenotypes, yet has a different causal gene and is ranked 16th. This capability of SHEPHERD to consider indirect, deep associations between genes and phenotypic features makes SHEPHERD highly complementary to graph theoretic techniques and statistical tests that can only score direct associations, which can be ineffective for poorly characterized diseases.

We next quantify SHEPHERD’s ability to identify “patients-like-me” for each UDN patient from all patients in the real-world MyGene2 cohort. As before, we evaluate the average recall at *k*, here defined as the number of MyGene2 patients with the same causal gene as the query correctly predicted in the top-*k* ranked patients on average for all UDN patients in the cohort. SHEPHERD ranks a patient with the same causal gene first in 11.5% of UDN patients, achieving a recall of 0.31, 0.43, 0.49, and 0.53 for *k* = 5, 10, 25, and 50, respectively (Figure 5a). We compare SHEPHERD to Phrank, an alternative approach that can calculate phenotypic similarity. Phrank uses information theory to calculate the similarity between two sets of phenotype terms based on shared ancestors in the Human Phenotype Ontology. We find that SHEPHERD performs significantly better than Phrank in identifying “patients-like-me” (Mann-Whitney *p*-value = 0.04). SHEPHERD ranks a patient with the same causal gene first for 7.4% more patients and reduces the number of patients that clinicians need to consider by 703 (17.2%) compared to Phrank.

Finally, we evaluate whether SHEPHERD embeds patients with the same disease (rather than gene) closer to each other than to patients with different diseases. Again, we compare UDN patients to MyGene2 patients. We find that embedding distances between patients diagnosed with the same disease are significantly smaller compared to patients with different diseases (*p*-value = 2.42 *×* 10^*−*8^; Kolmogorov-Smirnov test; Supplementary Figure 6), further strengthening the evidence that SHEPHERD can capture similarities between different diseases with similar presenting symptoms, but can nevertheless differentiate patients that have the same diagnosed disease.

### SHEPHERD provides an interpretable characterization of novel diseases

In addition to supporting causal gene discovery and patients-like-me identification, SHEPHERD can help characterize novel clinical presentations through our current knowledge of rare diseases (Figure 1a). Given a patient’s set of HPO phenotype terms, SHEPHERD provides an interpretable summary of the patient’s disease based on its similarity to each disease in the KG. SHEPHERD produces a ranked list of all diseases using the embedding similarity between each disease and the patient’s phenotype terms, which are then summarized to generate a distribution of similarities to disease categories. More concretely, SHEPHERD learns an embedding space in which the similarity between a patient and a disease is inversely proportional to the embedding distance between the patient and their diagnosed disease (Figure 6a). Aggregating SHEPHERD-generated similarities of individual diseases by their disease category enables interpretable characterization of the patient’s disease. For example, a patient’s presenting syndrome may be *w*_1_% similar to rare neurologic diseases, *w*_2_% similar to rare bone diseases, *w*_3_% similar to rare developmental defects during embryogenesis, etc. SHEPHERD can leverage gene-phenotype-disease associations to generate granular descriptions of a patient’s disease. For this analysis, we leverage patients from two cohorts: the simulated cohort is used for training, and the UDN cohort is used for validation.

**Figure 6:**
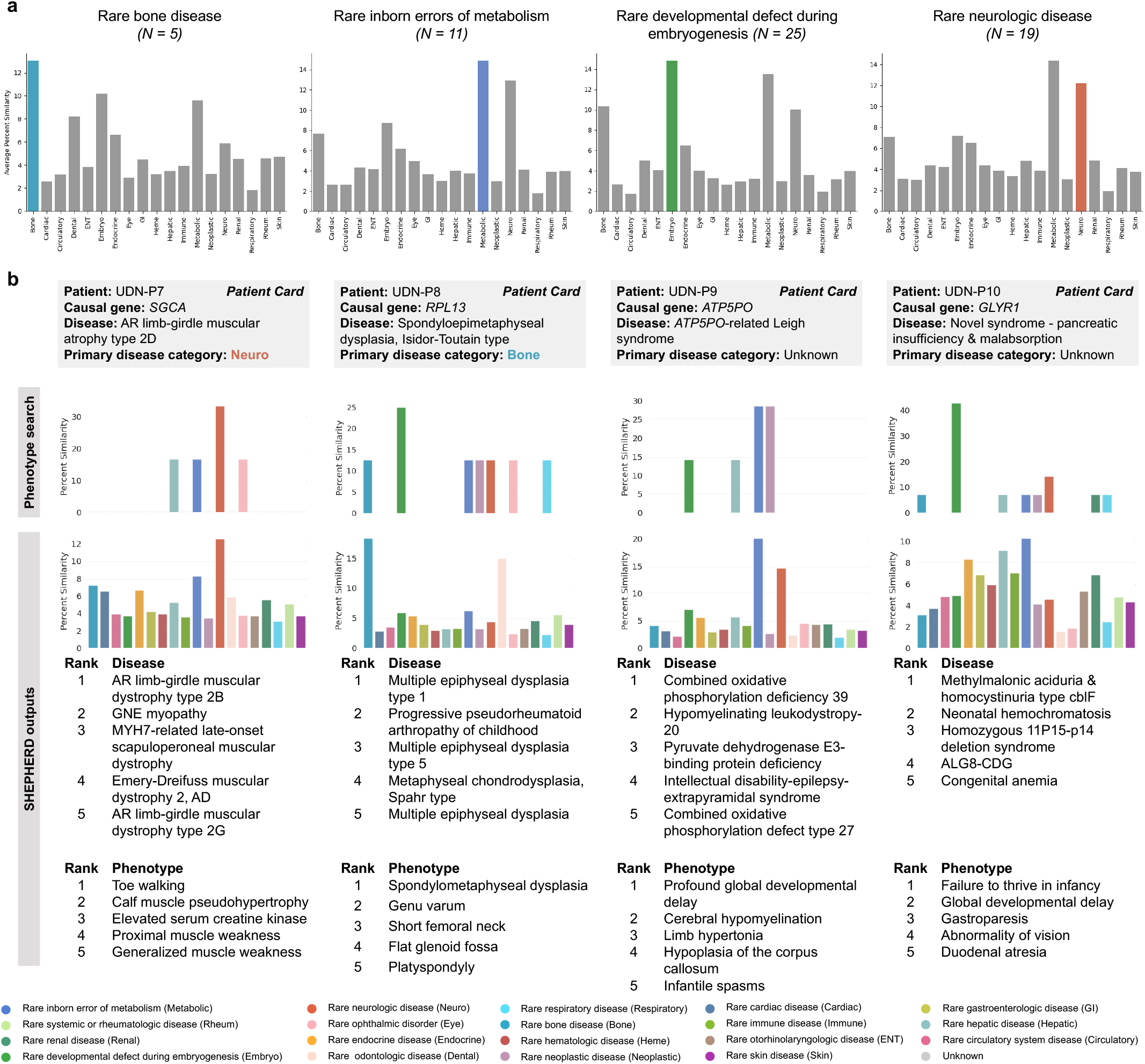
SHEPHERD performs novel disease characterization. **(a)** Bar plots of the similarity between UDN patients and diseases found in each disease category. We group UDN patients by the disease category of their true disease and show plots for all categories with at least 5 patients. The bars that do not correspond to the disease category of each patient’s true disease are colored gray. **(b)** The column for each of the four case studies contains: the percent similarity distributions of the patient’s phenotype terms to diseases in each disease category based on a phenotype search via the KG (top) or SHEPHERD (bottom), a table of the five most similar diseases according to SHEPHERD, and a table of the patient’s five phenotypic features that are most highly attended by SHEPHERD.

We observe that SHEPHERD learns to embed patients near diseases of the same category; on average, 45.7% of the top 10 ranked diseases with a known disease category belong to the same category as the patient’s disease, which is nearly three times more than random expectation alone (16.4%). To evaluate SHEPHERD’s ability to provide interpretable disease names for patients with known rare diseases, we first calculate the similarity between UDN patients and all diseases. This allows us to assess whether the patients are most similar to diseases that share the same disease category as the patient’s disease (Figure 6a). Concretely, for each patient, we stratify patients by their primary disease category and calculate the average similarity of a patient to all disease nodes under each disease category. As expected, we find that patients tend to be most similar to diseases of the same disease category as their own. For example, patients with a rare bone disease are predicted to be most similar to diseases under the category of rare bone disease (13.0% similarity), followed by rare developmental defects during embryogenesis (10.2%), rare inborn errors of metabolism (9.6%), and rare odontology diseases (8.2%). Similarly, patients with a disease categorized as a rare developmental defect during embryogenesis, a rare inborn error of metabolism, or a rare neurologic disease tend to be most similar to other diseases of the same category.

We examine two patients in depth to interrogate SHEPHERD’s predictive capabilities for characterizing known rare diseases: UDN-P7 and UDN-P8. Patient UDN-P7 (Figure 6b; SHEPHERD Tool Tab 3, Patient UDN-P7) received a diagnosis for limb-girdle muscular dystrophy 3 (sarcoglycanopathy; MIM:608099) due to variants in *SGCA*. SHEPHERD compares the patient’s clinical presentation to diseases across 19 disease categories and finds that the patient is most similar to rare neurologic diseases, as expected. From SHEPHERD’s predictions, two of the top five most similar diseases are other types of AR limb-girdle muscular dystrophy, and all five are related to muscular dystrophy. We compare SHEPHERD to a simple phenotypic search of the patient’s HPO terms to generate a distribution of similarities to disease categories. This phenotype search approach can correctly identify the patient’s disease as a rare neurologic disease but cannot produce disease-level rankings. Patient UDN-P8 (Figure 6b; SHEPHERD Tool Tab 3, Patient UDN-P8) was diagnosed four years after acceptance to the UDN with the bone disease spondyloepimetaphyseal dysplasia caused by a mutation in *RPL13*. Again, SHEPHERD can ascertain the patient’s symptoms are similar to other bone diseases; all of the top 5 ranked disorders are rare bone diseases with overlapping phenotype terms found in the query patient. In contrast, the phenotype search approach does not identify UDN-P8’s disease as a rare bone disease; rather, it predicts that the patient has a disease due to a rare developmental defect during embryogenesis. These findings on our case studies of two patients with known rare diseases suggest that SHEPHERD can produce correct and granular hypotheses about a patient’s rare disorder.

We also investigate SHEPHERD’s hypotheses for two patients with novel genetic diseases, UDN-P9 and UDN-P10. UDN-P9 (Figure 6b; SHEPHERD Tool Tab 3, Patient UDN-P9) was diagnosed with *ATP5PO*-related Leigh syndrome caused by a novel mutation in *ATP5PO*, a gene previously unassociated with any disease [61]. As Leigh syndrome is a metabolic disorder with neuropathological features, SHEPHERD correctly identifies UDN-P9’s disease as most similar to diseases under the categories of rare inborn errors of metabolism and rare neurological diseases. In contrast, the phenotype search method incorrectly predicts a tie between a disorder due to a rare inborn error of metabolism and a rare neoplastic disease, failing to label the patient’s disease as a neurological disorder. Three of the top five diseases—combined oxidative phosphorylation deficiency 39 (MIM:618397; ranked by SHEPHERD as #1), pyruvate dehydrogenase E3-binding protein deficiency (MIM:245349; ranked by SHEPHERD as #3), and combined oxidative phosphorylation defect type 26 (MIM:616672; ranked by SHEPHERD as #5)—are mitochondrial diseases affecting the same pathway as *ATP5PO* and result in a defect in the aerobic energy production. These diseases’ causal genes co-localize with *ATP5PO* [62–65]. Combined oxidative phosphorylation deficiency 39 and combined oxidative phosphorylation defect type 26 are associated with neurological presentations of mitochondrial disease, including hypotonia, seizures, and features of Leigh syndrome [66]. The remaining two most similar diseases (ranked by SHEPHERD as #2 and #4) are rare neurologic diseases with phenotype terms identical to UDN-P9’s. The causal gene, *CNP*, for the second-ranked disease, hypomyelinating leukodystropy-20 (MIM:619071), is three hops away from *ATP5PO* in the physical protein interaction network [67, 68], suggesting that they may be functionally related [69–71] or operate together [72, 73] to mediate phenotypic features associated with UDN-P9’s disease and hypomyelinating leukodystropy-20.

Patient UDN-P10 (Figure 6b; SHEPHERD Tool Tab 3, Patient UDN-P10), is characterized by SHEPHERD as most similar to diseases under the categories of rare inborn errors of metabolism, rare hepatic disease, rare gastroenterological disease, and rare endocrine disease. These top categories are aligned with many of the patient’s symptoms, particularly duodenal atresia, intestinal malrotation, pancreatic exocrine insufficiency, liver disease, and developmental delay. In contrast, the phenotype search approach predicts that the patient’s disease is most similar to diseases due to rare developmental defects during embryogenesis. Three of the top five most similar individual diseases from SHEPHERD’s outputs—Methylmalonic acidemia with homocystinuria type cblF (MIM:277380; ranked by SHEPHERD as #1), Neonatal hemochromatosis (MIM:231100; ranked by SHEPHERD as #2), and ALG8-CDG (MIM:608104; ranked by SHEPHERD as #4)—are also due to inborn errors of metabolism, and the diseases are associated with phenotypes that are similar to those seen in the patient, including abnormalities in liver and gastrointestinal function and developmental delay. Notably, the rare respiratory disease category is the third lowest-ranked category. UDN clinicians hypothesized that the patient’s *GLYR1* variants cause a mislocalization of the cystic fibrosis conductance regulator (*CFTR*), which is associated with cystic fibrosis. While the patient has gastrointestinal and pancreatic symptoms similar to those in cystic fibrosis, the patient does not have any of the pulmonary features classic for that condition. Such granularity in SHEPHERD’s predictions is a reflection of SHEPHERD’s ability to differentiate between diseases despite partially overlapping phenotypes and causal genes sharing the same pathway.

## Discussion

We present SHEPHERD, a deep learning approach for multi-faceted rare disease diagnosis. SHEPHERD overcomes limitations of supervised deep learning by (1) incorporating biomedical knowledge into the model via geometric deep learning on a knowledge graph, (2) leveraging label-efficient learning to align patients with genes and phenotypes, and (3) training on a large dataset of simulated rare disease patients in a disease-stratified manner. Further, the attention weights that are learned for generating phenotype-based patient embeddings can be inspected to provide insights into the contribution of each phenotype term to the patient-specific prediction. As shown in the evaluations on external multi-site patient cohorts with heterogeneous disease presentations, SHEPHERD generalizes across phenotype terms, genes, and diseases and performs well on patients with heterogeneous clinical presentations and novel genetic conditions (Supplementary Figure 2). A unique feature of SHEPHERD is its ability to generate clinico-genetic representations of patients with rare genetic diseases. SHEPHERD represents patient phenotype terms as subgraphs and candidate genes and diseases as nodes in the knowledge graph. The graph neural network then generates embeddings by considering direct and indirect gene-phenotype-disease associations that are multiple hops away from each other in the knowledge graph. While many existing approaches rely exclusively on known phenotype-gene-disease associations [16, 18], leveraging indirect associations is essential for diagnosing patients with novel or atypical genetic conditions (Supplementary Figure 3b, Supplementary Figure 4d). Further, subgraphs provide a flexible mathematical definition for modeling sets of the patient phenotype terms. Rather than modeling each phenotype term individually [19], SHEPHERD encodes patients as a structured object (*i*.*e*. a subgraph) and considers the co-occurrence of phenotypic features when diagnosing rare diseases.This joint modeling of patient phenotypic features is essential for identifying genetic mutations with pleiotropic effects [74–76]. This approach also helps mitigate variability in phenotype annotations by leveraging relationships in the knowledge graph to connect patients described with different but related phenotype terms.

SHEPHERD demonstrates that models trained on simulated patient datasets apply to realworld clinical applications. While simulated data is increasingly used to augment training datasets for improving robustness and generalizability [52,77–81], here we primarily use simulated patients to train SHEPHERD. Simulated data are not just an additional asset, but a critical necessity for training deep learning models to generate predictions on rare diseases with scarce labeled diagnoses. The synthetic patients are generated by a simulator [48] based on clinico-genetic knowledge. Training on simulated data mitigates concerns regarding privacy breaches, in which specific individuals can be identified from the training data [82, 83]. Hence, a fully trained SHEPHERD model can be publicly released without privacy concerns.

There are several extensions to this work. Our method relies on a knowledge graph of disease, gene, and phenotype associations. Other sources of information, such as variant-level information or databases of model organism phenotype-gene associations, can be incorporated as well [84]. SHEPHERD’s knowledge graph includes gene-phenotype-disease associations and can be extended to include information from research literature [26]. SHEPHERD’s phenotype-based approach can also be combined with variant-based prioritization approaches, such as those used in Exomiser [24], for even stronger causal gene discovery performance. The graph neural network underlying SHEPHERD can be extensible to multi-modal data types. For example, gene co-expression data or textual descriptions of diseases can be incorporated as node features. Given the importance of negative phenotypes (*i*.*e*., explicitly absent symptoms) for differential diagnosis, extending SHEPHERD to consider both present and absent symptoms may improve patient representations for multi-faceted rare disease diagnosis. Incorporating temporal modeling of phenotype progression into the patient simulation process and SHEPHERD’s framework could further enhance its ability to recognize age-dependent disease manifestations and improve diagnostic accuracy across the lifespan. It is also worth exploring various graph transformer architectures to potentially improve SHEPHERD’s ability to harness the benefits of global attention while preserving the graph’s structural nuances. Recent large language model (LLM) approaches for rare diseases could complement SHEPHERD by leveraging biomedical pretraining and retrieval-augmented generation to support diagnosis from unstructured clinical notes and literature without requiring structured phenotype terms [85–87] However, out-of-the-box LLMs have been shown to underperform in rare disease diagnosis [88], likely due to the rarity of many conditions in biomedical corpora. Hybrid models that either incorporate graph-based representations into LLMs or integrate LLM-derived embeddings into structured knowledge graphs may help mitigate these limitations, enhancing diagnostic accuracy and interpretability. Evaluating the utility of SHEPHERD in prospective clinical workflows represents an important next step to assess its performance in practice and understand its impact on clinical decision-making. Finally, while efforts like the UDN are critical for establishing diagnoses for rare disease patients, they alone cannot address the rare disease burden. Approaches like SHEPHERD can help identify and diagnose rare disease patients using claims data, electronic health records, and other data types. SHEPHERD’s ability to characterize a patient’s clinical presentation can be used to identify sub-specialists who should review the patient’s case for the diagnostic recommendation.

Our study has a few limitations. First, continually updating the knowledge graph with genephenotype-disease associations can improve SHEPHERD’s performance. To this end, the knowledge graph curation and processing approaches are fully reproducible, and the graph can be automatically updated as new data become available [89]. Second, the still-undiagnosed UDN patients may be more challenging than the already-diagnosed ones SHEPHERD was tested on. There are two categories of still-undiagnosed patients: patients admitted to the UDN years ago who have yet to receive a diagnosis due to sequencing limitations (*e*.*g*., hard-to-detect variant types such as short tandem repeats or structural variants, missing second variants in recessive disorders, variants that lie in difficult-to-sequence regions or are masked due to biases in the human reference genome and ancestral genomes [90]), and patients recently admitted to the UDN. SHEPHERD can be evaluated on the still-undiagnosed patients whose causal variants will be detectable by deep whole genome sequencing. The lack of an observed drop in SHEPHERD’s performance for recently diagnosed patients suggests that data leakage (*i*.*e*., information about older diagnoses being incorporated into the knowledge graph) has not occurred, evidently avoiding the bias that would otherwise cause overfitting of the model to the training data. Finally, like many genomic studies, our datasets likely overrepresent individuals of European descent. This demographic bias is an important limitation that could affect generalizability, and highlights the need for more inclusive rare disease datasets in future work.

SHEPHERD shows the utility and impact of deep learning for diagnosing rare disease patients. While other deep learning-powered diagnostic systems focus on common diseases for which large labeled datasets exist, this study shows how deep learning can be used for rare diseases. The diagnostic process requires collaborations among bioinformaticians, clinicians, and genetic counselors. Reviewing a single case can take many hours of a many-person team over days or weeks. SHEPHERD can substantially reduce the number of genes that human experts need to consider to provide a molecular diagnosis and identify patients with similar genetic conditions, even before they have undergone genetic sequencing. Deep learning-based diagnostic strategies like SHEPHERD create new opportunities to shorten the diagnostic odyssey for rare diseases.

## Methods

The Methods section is structured as follows: description of our rare disease knowledge graph; details of our rare disease patient cohorts; formulations of SHEPHERD, our algorithmic approach for rare disease diagnosis; details regarding model training; and descriptions of our statistical analysis and evaluation setup.

### 1 Rare Disease Knowledge Graph Construction

We create a comprehensive knowledge graph (KG) for rare disease diagnosis. We start with PrimeKG [89] and adapt it to the rare disease diagnostic setting by removing drug-specific entities and relations and incorporating additional sources of the gene, phenotype, and disease relationships. The resulting rare disease KG contains seven node types (*i*.*e*., phenotype, protein, disease, pathway, molecular function (MF), cellular component (CC), and biological process (BP)) and 15 unique relation types (*i*.*e*., phenotype-protein, disease-phenotype^(-)^ (indicating that disease does not have phenotype), disease-phenotype^(+)^ (indicating that disease has phenotype), protein-pathway, disease-protein, protein-MF, protein-CC, protein-BP, BP-BP, MF-MF, CC-CC, phenotype-phenotype, protein-protein, disease-disease, pathway-pathway).

Relationships are extracted from the following data sources: Gene Ontology (GO) [96], Reactome pathway knowledgebase [95], DisGeNET [98], NCBI [102], Human Phenotype Ontology (HPO) [92], MONDO disease ontology [93], and Orphanet [97]. PrimeKG contains disease-protein relationships from DisGeNET, and we include additional disease-protein and disease-phenotype relationships from Orphanet if they are not already present in the KG. All phenotype terms are mapped to the Human Phenotype Ontology, all genes/proteins are mapped to Ensembl identifiers, and all diseases are mapped to MONDO identifiers. When a concept is represented in the HPO and MONDO ontologies, we remove the MONDO identifier. This differs from the original PrimeKG preprocessing, where conflicting identifiers are mapped to MONDO IDs. We perform all other preprocessing as in the original PrimeKG knowledge graph. In particular, duplicate and self-loop edges are removed, and only the largest connected component of the graph is retained to ensure connectivity. For additional information about each data source and the harmonization process, refer to PrimeKG [89].

We enforce homophily between genes and phenotypes by computing the triadic closure between gene-disease and disease-phenotype edges [103, 104]. We extract the largest connected component to ensure the KG is fully connected. The largest connected component retains 99.91% of the nodes and 99.99% of the edges from the knowledge graph. Finally, we add reverse edges to ensure the KG is represented as an undirected graph during model training.

The final knowledge graph contains 105,220 nodes and 1,095,469 edges. Tables 1-2 outline the number of nodes and edges by node type and relation type, respectively.

**Table 1:** Statistics about nodes in the rare disease knowledge graph. Reported is the number of nodes by node type. MF: molecular function, CC: cellular component, BP: biological process.

| NODE TYPE | COUNT | AVERAGE DEGREE | VOCABULARY |
| --- | --- | --- | --- |
| PHENOTYPE | 15,874 | $49.5 \pm 190.6$ | HUMAN PHENOTYPE ONTOLOGY [92] |
| DISEASE | 21,233 | $17.1 \pm 37.4$ | MONDO [93] |
| PROTEIN | 21,610 | $74.2 \pm 119.8$ | ENSEMBL [94] |
| PATHWAY | 2,516 | $19.0 \pm 29.9$ | REACTOME [95] |
| MF | 11,169 | $8.7 \pm 127.4$ | GENE ONTOLOGY [96] |
| CC | 4,176 | $22.3 \pm 197.2$ | GENE ONTOLOGY [96] |
| BP | 28,642 | $8.7 \pm 28.1$ | GENE ONTOLOGY [96] |

**Table 2:** Statistics about the edges in the rare disease knowledge graph. Reported is the number of edges by relation type. HPO: Human Phenotype Ontology.

| RELATION TYPE | COUNT | SOURCES |
| --- | --- | --- |
| PHENOTYPE-PHENOTYPE | 21,925 | HPO [92] |
| PHENOTYPE-PROTEIN | 10,518 | HPO [92], MONDO [93], ORPHANET [97] |
| DISEASE-DISEASE | 35,167 | DISGENET [98], MONDO [93], ORPHANET [97] |
| DISEASE-PHENOTYPE<sup>(-)</sup> | 1,483 | HPO [92], DISGENET [98], MONDO [93], ORPHANET [97] |
| DISEASE-PHENOTYPE<sup>(+)</sup> | 204,779 | HPO [92], DISGENET [98], MONDO [93], ORPHANET [97] |
| DISEASE-PROTEIN | 86,299 | DISGENET [98], MONDO [93], ORPHANET [97] |
| PROTEIN-PROTEIN | 321,075 | MENCHE *et al.* [99], BIOGRID [100], STRING [101], LUCK *et al.* [73] |
| PROTEIN-PATHWAY | 42,646 | REACTOME [95] |
| PATHWAY-PATHWAY | 2,535 | REACTOME [95] |
| PROTEIN-MF | 69,530 | GENE ONTOLOGY [96] |
| PROTEIN-CC | 83,402 | GENE ONTOLOGY [96] |
| PROTEIN-BP | 144,805 | GENE ONTOLOGY [96] |
| MF-MF | 13,574 | GENE ONTOLOGY [96] |
| CC-CC | 4,845 | GENE ONTOLOGY [96] |
| BP-BP | 52,886 | GENE ONTOLOGY [96] |

### 2 Rare Disease Patient Cohorts

We use four distinct rare disease patient cohorts for training and evaluating SHEPHERD: UDN (Section 3), a real-world cohort of hard-to-diagnose patients in the Undiagnosed Diseases Network, MYGENE2 (Section 4), a publicly available real-world cohort of patients with rare genetic conditions who have opted to share their information on the MyGene2 Portal, DDD (Section 5), a publicly available aggregated summary of real pediatric patients with severe developmental disorders in the Deciphering Developmental Disorders study, and SIMULATED (Section 6), a large diverse and realistic simulated patient cohort representing 2,134 unique rare diseases in Orphanet. The diseases found in each cohort are in Supplementary Table 1.

For every patient cohort, we categorize each patient’s causal disease according to the 33 disease categories outlined in Orphanet. We map all diseases to Orphanet and leverage the Orphanet linearisation process (http://www.orphadata.org/cgi-bin/rarefree.html) to assign each disease to a single disease category based on a series of rules that consider the most severely affected body system and the specialists most likely to be involved in treatment.

### 3 Patients in the Undiagnosed Diseases Network

The Undiagnosed Diseases Network (UDN) is a nationwide research study supported by the National Institutes of Health Common Fund, which aims to bring together clinical and research experts around the United States to diagnose patients with rare genetic conditions [105]. The Undiagnosed Diseases Network study is approved by the National Institutes of Health institutional review board (IRB), which serves as the central IRB for the study **(IRB Protocol 15HG0130)**. All patients accepted to the UDN provide written informed consent to share their data across the UDN as part of a network-wide informed consent process.

The UDN consists of 12 clinical sites across the United States that evaluate patients, a sequencing core, a model organism screening center, a central biorepository, a metabolomics core, and a coordinating center. Patients are admitted to the UDN if they have objective findings, and clinical testing has failed to produce a diagnosis. Most admitted patients receive exome or whole genome sequencing and an extensive clinical workup.

We include patients from the Undiagnosed Diseases Network who meet the following criteria: (1) at least one phenotype term describing their clinical presentation, (2) at least five candidate genes potentially explaining their symptoms, and (3) a diagnosis classified as ‘Certain’ or ‘Highly Likely’ based on the UDN’s diagnostic certainty annotations [29]. Diagnoses with minimal uncertainty are considered ‘Certain,’ while those with some uncertainty—yet still sufficient for clinical decision-making—are classified as ‘Highly Likely.’

We construct patient subgraphs using the phenotypes obtained through deep phenotyping. Deep phenotyping of patients during the clinical workup is a central component of the UDN process. Clinicians annotate each patient with a set of terms from the Human Phenotype Ontology (HPO) using PhenoTips, a tool integrated into the electronic health record that allows for structured phenotyping of patient symptoms [106]. We map all phenotype terms to the same version of the Human Phenotype Ontology (v2019), discard 406 unique prenatal phenotype terms related to the mother’s pregnancy and use all remaining phenotype terms to construct patient subgraphs. Each patient subgraph is formed from the phenotype nodes in the rare disease knowledge graph that describe the patient’s symptoms (Methods 8). We construct phenotype subgraphs for the 465 UDN patients with annotated phenotype terms who have received a molecular diagnosis as of January 5, 2022.

We obtain EXPERT-CURATED candidate gene lists from the UDN. Genomic samples for each patient are sequenced at Baylor Genetics or Hudson Alpha. All candidate genes are standardized to Ensembl gene identifiers. We construct an EXPERT-CURATED candidate gene list for each patient from the patient’s sequencing data. Importantly, these gene lists are unique to each patient. The EXPERT-CURATED candidate gene list for each patient includes the union of both (1) disease-associated and other clinically-relevant genes listed on the patient’s clinical sequencing reports from Baylor or Hudson Alpha per the UDN protocol and American College of Medical Genetics and Genomics (ACMG) guidelines [29, 107, 108] and (2) genes that were prioritized by UDN clinical teams who handled the patient’s case. The genes in this list represent the strongest candidates identified by the UDN sequencing core or the clinical team. In addition, the list often includes known disease-causing genes, genes with suspected pathogenic variants, or genes expressed in tissues relevant to the patient’s clinical presentation. While the EXPERT-CURATED gene list contains the strongest candidates, the list nevertheless requires further filtering to identify the ultimate causal gene(s) that explain the patient’s condition. We exclude patients whose candidate gene lists have fewer than five candidate genes for the causal gene discovery task. The cohort contains 278 patients with at least five EXPERT-CURATED candidate genes.

We obtain VARIANT-FILTERED candidate gene lists from the UDN. As part of the UDN analysis pipeline, the UDN performs the whole genome and exome sequencing for a patient and their immediate family members. Here, we use the patients’ whole genome sequencing (WGS) data, which are aligned to the GRCh38.p13/hg38 human genome build and have undergone variant calling via the Genome Analysis Toolkit (GATK) best practices workflow [50]. Please refer to [50] for more details about the computational workflow across UDN sites. Access to the UDN patients’ WGS data allows us to construct for each patient a VARIANT-FILTERED candidate gene list consisting of genes that have at least one variant and that have been prioritized by a variant-level prioritization algorithm. We leverage the Exomiser algorithm, which considers variant frequency, predicted pathogenicity, and (if family members’ sequencing data are available) mode of inheritance [109]. While Exomiser can leverage known associations between genes and phenotypic features, we do not use it to construct our VARIANT-FILTERED candidate gene lists. We analyze the patients’ variant-called WGS data (*i*.*e*. variant call format, or VCF) using Exomiser under the following inheritance modes: autosomal dominant, autosomal recessive homozygous alternate, autosomal recessive compound heterozygous, X dominant, X recessive homozygous alternative, X recessive compound heterozygous, and mitochondrial. Their respective cutoff values (*i*.*e*. the maximum minor allele frequency in percent (%) permitted for an allele to be considered a causative candidate under that mode of inheritance) are 0.1, 0.1, 2.0, 0.1, 0.1, 2.0, and 0.2. We remove variants with non-coding effects (*i*.*e*. 5’ and 3’ UTR exon/intron variants, non-coding transcript exon/intron variants, coding transcript intron variants, up-/down-stream gene variants, intergenic variants, and regulatory region variants). We use the following pathogenicity sources, `POLYPHEN`, `MUTATION TASTER`, and `SIFT`. We apply a frequency filter to remove variants with a frequency of at least 0.5% according to the variant frequency databases used. All variant frequency databases are used, as recommended by the Exomiser manual. We retain non-pathogenic variants in the output gene list. As with the EXPERT-CURATED gene lists, we filter out patients who do not have at least five candidate genes in their VARIANT-FILTERED gene list. The cohort includes 229 patients with at least five VARIANT-FILTERED candidate genes.

Diagnosed patients in the UDN are labeled with a disease identifier from the Online Mendelian Inheritance in Man (OMIM) database [110] when the patient is diagnosed with a known genetic disease. We map the OMIM disease identifiers to MONDO identifiers [93] using the MONDO ontology crosswalk to identify the diseases in the rare disease knowledge graph (Section 1).

The final UDN cohort contains 465 patients representing 319 MONDO diseases and 378 unique causal genes. The EXPERT-CURATED and VARIANT-FILTERED candidate gene lists contain 244.3 and 13.3 genes on average, respectively (SD = 244.0 and SD = 8.0). Patients have 23.9 HPO phenotype terms on average (SD = 16.1).

### 4 Patients in the MyGene2 Portal

We assemble a cohort of real-world rare disease patients participating in the MyGene2 exchange [53]. MyGene2, developed by the University of Washington, is a portal through which families with rare genetic conditions can share their health information to connect with other families, clinicians, and researchers. MyGene2 contains information about 2,106 genes and the HPO phenotype terms of patients with gene mutations. MyGene2 is a member of the MatchMaker Exchange, a federated network designed to enable clinicians to find phenotype and genotype matches for rare disease patients [111]. The UDN leverages the MatchMaker exchange to validate patients’ candidate genes by finding genotype-matched individuals.

We retrieved data containing the sets of phenotype terms and candidate genes for rare disease patients on MyGene2 as of May 7, 2022. We filter the patients only to include patients labeled with an OMIM disease identifier and a single candidate gene. This limits the cohort to patients who are likely already diagnosed. As with the other cohorts, we map all genes to Ensembl identifiers and diseases to MONDO identifiers, and construct patient subgraphs from the set of positive HPO terms associated with each patient. Demographic information, such as age, sex, or ancestry, is not systematically collected in MyGene2 and is therefore unavailable for this cohort.

The final MyGene2 cohort contains 146 patients representing 55 MONDO diseases and 48 unique causal genes. Patients have 7.9 HPO phenotype terms on average (SD = 6.6). There are 14 unique causal genes and 12 diseases in the MyGene2 and UDN cohorts.

### 5 Patients derived from the Deciphering Developmental Disorders Study

We construct another dataset of rare disease patients using aggregated gene and phenotypic information from patients in the Deciphering Developmental Disorders Study. This initiative recruited nearly 14,000 children with severe undiagnosed developmental disorders from the United Kingdom and Ireland [54]. Among enrolled probands, 42% were female, 16% were of non-European ancestry, and the median age at recruitment was 7 years.

We retrieved data containing the sets of phenotype terms and associated genes from DECIPHER (https://www.deciphergenomics.org/ddd/ddgenes) on May 10, 2023. We remove genes where the evidence supporting a causal role for the gene is either limited or moderate, and we use the remaining genes and associated phenotype term sets to construct the patient cohort. We map all genes to Ensembl identifiers and diseases to MONDO identifiers, and build patient subgraphs from the set of HPO terms associated with each patient. Non-causal candidate genes for each patient are constructed by sampling genes in the knowledge graph neighboring the patient’s phenotype terms or causal gene.

The final DDD-derived cohort contains 1,431 patients, representing 1,237 MONDO diseases and 1,282 unique causal genes. Patients have 20.5 HPO phenotype terms on average (SD = 19.2). There are 158 unique causal genes and 93 diseases found in the DDD and UDN cohorts.

### 6 Simulated Patients with Rare Mendelian Disorders

We leverage simulated but realistic rare disease patients for training SHEPHERD [48]. The simulated patients closely resemble real-world patients found in the UDN. Each simulated patient is represented by an age range, a set of positive phenotypic features they exhibit, a set of negative phenotypic features they do not exhibit, and a set of challenging candidate genes that may cause the presenting symptoms. The patients are generated using a simulation framework that jointly samples candidate genes and phenotype terms.

To generate patients with rare Mendelian disorders, we adopt the pipeline described in [48]. Briefly, the simulation pipeline has two stages: phenotype and candidate gene generation. First, each patient is initialized with a set of phenotype terms associated with a genetic disorder characterized in the rare genetic disease database Orphanet [97]. To reflect the imprecision of real-world diagnostic evaluations, the initial phenotype terms undergo phenotype dropout and corruption (*i*.*e*., phenotype terms are randomly removed or replaced with more general phenotype terms), and additional “noisy” phenotype terms that are unrelated to the patient’s disease are sampled from a large medical insurance claims database and added to the phenotype set. Next, candidate genes are sampled from “distractor” gene categories that do not cause the patient’s disease yet would be plausible candidates during the diagnostic process. The challenging distractor genes and some of their associated phenotype terms are added. For additional details about the simulation process and validation of simulated patients, refer to [48]. To standardize across all patient cohorts, we ensure all genes are mapped to Ensembl identifiers, all diseases are mapped to MONDO identifiers, and we construct patient subgraphs from the phenotype terms associated with each patient.

There are 42,624 simulated patients representing 2,132 unique Mendelian disorders and 2,396 unique causal genes in the simulated patient dataset. Each patient is characterized by an average of 18.4 positive phenotype terms (SD = 7.7) and 14.0 candidate genes (SD = 3.5). Of the 378 unique causal genes and 319 unique MONDO diseases found in patients in the UDN cohort, 220 and 109 are represented in the simulated patient cohort, respectively. Furthermore, 81.8% of the phenotype terms found across UDN patients are also found in the simulated patient cohort, and 29.7% of a single UDN patient’s phenotype terms are found in the most similar simulated patient on average. This indicates that the simulated patients have utility in training models that can apply to real-world UDN patients but also emphasizes the need for developing models that can generalize to genes, diseases, and phenotype terms unseen at train time.

### 7 Few shot Learning Framework for Rare Disease Diagnosis

We develop SHEPHERD, a geometric deep learning approach that leverages few shot capability and external biomedical knowledge for multi-faceted diagnosis of rare diseases. SHEPHERD learns to co-embed diseases, phenotypes, and genes for generating multi-modal representations of rare disease patients. As such, it performs multi-faceted diagnosis, addressing the following challenges of rare disease diagnosis: Causal gene discovery, identification of similar patients, and characterization of novel diseases.

For **causal gene discovery**, each patient *T*_*i*_ in the dataset has *P*_*i*_ phenotype terms and *G*_*i*_ candidate genes. The task is to identify the causal gene(s) 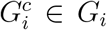, harboring the variants that explain the patient’s presenting symptoms.

For the **identification of similar patients**: Given a cohort of rare disease patients *C*, the goal is to identify patients from the cohort similar to the query patient *T*_*i*_ (*i*.*e*., patients that share a disease or causal gene). Mathematically, for each patient *T*_*i*_, the task is to identify the set of Patients 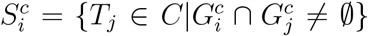. We leverage each patient’s set of phenotype terms *P*_*i*_ to perform patient matching.

For the **characterization of novel diseases**, the goal is to characterize novel diseases according to their similarity to a set of known genetic diseases *D*. We input the set of phenotype terms *P*_*i*_ for each patient *T*_*i*_ and provide interpretable names for the patient’s presenting syndrome.

### 8 Notation

Let *𝒢* denote a heterogeneous knowledge graph comprised of a set of nodes *V* and a set of edges *E*. Each edge is defined by a triplet (*u, r, v*) where *u* is the source node, *v* is the target node and *r ∈ R* denotes the relationship between *u* and *v*. Each patient *i* is represented on the graph as a patient subgraph induced by a set of phenotype nodes *P*_*i*_ where *P*_*i*_ *⊆ V*. The patient subgraph can contain any number of phenotype terms and multiple connected components throughout *𝒢*. Each patient may also have a set of candidate genes *G*_*i*_ *⊆ V*.

### 9 SHEPHERD: Encoding Biomedical Knowledge

The first step in SHEPHERD is to encode biomedical relationships in the rare disease knowledge graph (KG). Here, we describe the architecture of SHEPHERD’s GNN encoder, and then the objective function for pretraining SHEPHERD.

We begin by describing SHEPHERD’s GNN encoder **architecture**. We pretrain SHEPHERD on millions of biomedical entity pairs across all entity and relation types in the KG to capture the topology of the KG. To this end, we use a graph attention network (GAT) [112], a type of graph neural network (GNN) model to generate embeddings **x**_*v*_ for every node *v* in the KG. Specifically, the choice of a graph attention network is necessary to achieve semantically-relevant mixing of biomedical entities in the embedding space; that is, to encourage distinct node types (*e*.*g*. genes, diseases, and phenotypes) to be positioned near each other in the embedding space. Like most GNNs, GAT models can be formulated as message-passing networks, in which messages are propagated to a node *v* from all of the nodes in its neighborhood *𝒩* _*v*_. The messages are aggregated and combined with the previous layer’s representation of *v* to produce *v*’s representation for the current layer. Concretely, each layer *l* in SHEPHERD’s GNN encoder involves the following steps:

The first step is to **propagate neural messages**. We define the message 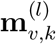 for each node *v* as:

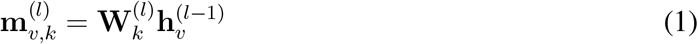

where *k* represents the attention head, **W** is a trainable weight matrix, and **h**_*v*_ is the embedding of node *v* in the (*l −* 1)-th hidden layer.

The second step is to **aggregate messages from local neighborhoods**. We leverage the local neighborhood to generate a representation of each node *v*. Specifically, we aggregate messages of its neighboring nodes *u ∈𝒩*_*v*_ and itself using an attention mechanism to generate 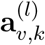:

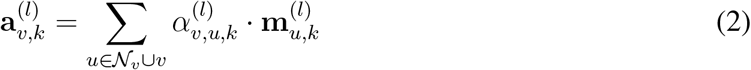

where *α*_*v,u,k*_ is the normalized attention weight on an edge from node *v* to node *u* computed by the *k*-th attention mechanism.

The third step is to **update node embeddings**. To transform the messages into an orderinvariant hidden representation 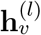, we apply a nonlinearity function *σ* and concatenate all of the aggregated messages as follows:

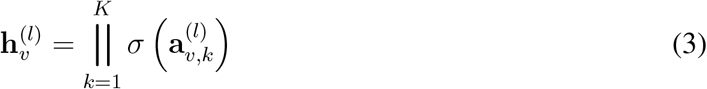

In the final layer, we perform averaging instead of concatenation. We define the final embedding for each node *v* after *L* layers of neural message passing as 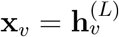. We specify *L* = 3 layers of neural message passing.

Next, we define SHEPHERD’s pretraining **objective function**. We frame pretraining as a binary classification task. SHEPHERD learns to perform link prediction (*i*.*e*. predict whether a relationship exists between a pair of nodes for a given relation type). Formally, we compute the score for whether an edge exists between node *u* and node *v* with relation *r* given their node embeddings **x**_*u*_ and **x**_*v*_ using a DistMult decoder [113]:

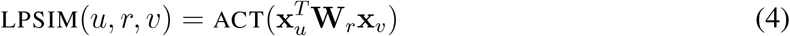

where **W**_*r*_ is a relation-specific trainable weight matrix and ACT is a nonlinear function (here, tanh). SHEPHERD is pretrained via a hinge loss objective. For any pair of nodes *u* and *v* connected by relation *r*, the loss function is defined as:

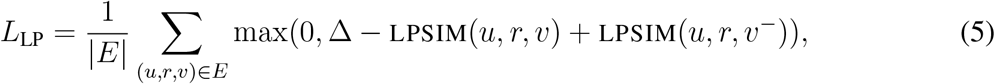

where *u* and *v* are source and target nodes, *v*^*−*^ is a target node representing a negative example that is not linked to *u* in the KG, LPSIM returns the score indicative of the knowledge relationship existing between *u* and *v*, and Δ is a margin, which is set to 1 throughout all experiments in this study. For each triplet, (*u, r, v*) in the KG, its contribution to the value of the loss function is 0 if the difference between the LPSIM’s score for the triplet and the LPSIM’s score for a negative example is at least as large as the margin.

### 10 SHEPHERD: Generating Rare Disease Patient Representations

We apply the pretrained SHEPHERD model to our multi-faceted rare disease diagnosis tasks. Starting with the pretrained GNN model, we learn patient embeddings that encode each patient’s phenotype subgraph. Depending on the diagnostic task, we also learn embeddings for each patient’s candidate genes, diseases, or other patients. Concretely, for every patient *T*_*i*_, we generate an aggregated representation of all phenotype terms *p ∈ P*_*i*_ in the phenotype subgraph via a transformer encoder and an attention-weighted average of the individual phenotype embeddings:

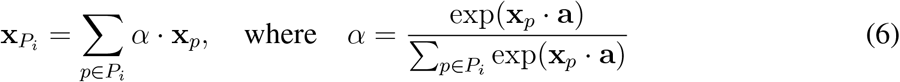

where *α* denotes the attention weights, **x**_*p*_ denotes the embedding for phenotype term *p*, and **a** is a trainable vector initialized via Xavier [114]. The aggregated phenotype representation 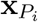, each candidate gene node embedding **x**_*g*_, and each candidate disease node embedding **x**_*d*_ are pushed through two nonlinear layers to produce the embeddings **z**_*P*_*i*, **z**_*g*_, and **z**_*d*_, respectively, as:

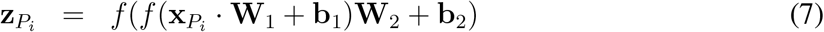

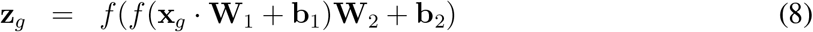

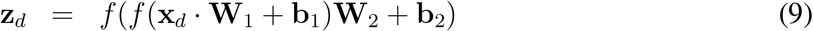

where *f* is a nonlinear function (here, leaky ReLU), and **W**_1_, **W**_2_, **b**_1_, and **b**_2_ are trainable weights. The embeddings, 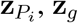, and **z**_*d*_, are each *m*-dimensional, where the output dimension *m* is determined via hyperparameter search (Section 18).

Finally, each candidate gene’s embedding **z**_*g*_ may be augmented with the embeddings of the *K* most similar genes. First, an aggregated embedding 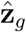 of the *K* genes with the highest number of shared phenotype terms as gene *g* is generated:

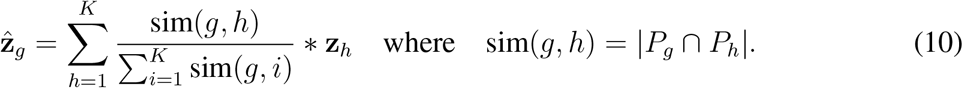

The original gene embedding is then updated via a gating mechanism as follows:

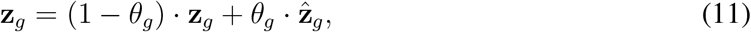

where *θ*_*g*_ controls the contribution of the original gene embedding for gene *g*. We set *θ*_*g*_ = *ω \**exp(*−ω \** | *𝒩* _*g*_|) + 0.2 where *ω* is a hyperparameter for the contribution of the augmented gene embedding, and | *𝒩* _*g*_| is the node degree for gene *g* to preferentially update the embeddings for genes with lower degree (*i*.*e*., genes that are not as well-characterized) [115].

This approach is motivated by the observation that novel disease genes (*i*.*e*., genes without known associations with any disease) or diseases (*i*.*e*., diseases without known associations with any gene) can have limited prior research or understanding, resulting in scarce neighbors in the knowledge graph. Due to this sparsity, the gene nodes’ embeddings are of lower quality, which can negatively impact the ability to identify the causal gene of a patient. For example, in an extreme case, a gene node without any connections to the rest of the knowledge graph would have a randomly initialized embedding. As such, we leverage information about shared phenotypic neighbors to augment these low-information gene nodes.

### 11 SHEPHERD: Discovering Causal Genes

SHEPHERD can prioritize candidate genes to assist clinicians in finding the causal gene(s) harboring the variants that best explain a patient’s presenting symptoms. Candidate genes for each patient are scored by measuring the similarity SIM(*P, g*) between a candidate gene *g* and a patient’s set of phenotype terms *P*. SHEPHERD is optimized such that the candidate gene embedded near the patient’s set of phenotype terms in the embedding space indicates that the gene will likely cause the patient’s symptoms. SIM(*P, g*) consists of two components, EMBSIM(*P, g*) and SPLSIM(*P, g*). It is designed such that EMBSIM(*P, g*) captures global network topology (*i*.*e*., by leveraging SHEPHERD’s low-dimensional embedding space) and SPLSIM(*P, g*) captures local network information (*i*.*e*., by calculating shortest path length distances). This approach is grounded in the observation that, while methods that learn global network topology yield higher overall performance than local methods considering only local network information, the latter tends to rank true candidate genes higher when provided a short list of candidate genes [116].

We calculate EMBSIM, an **embedding-based similarity** between aggregated embeddings of phenotype terms *P* and an embedding of the candidate gene *g* as follows:

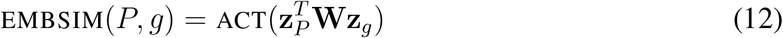

where ACT is a nonlinear function (here, tanh). EMBSIM values range between [*−*1, 1]. Analogous to LPSIM (*i*.*e*., self-supervised link prediction), EMBSIM predicts whether there exists a relationship (*i*.*e*., harbors variants that explain presenting symptoms) between a patient and the candidate gene. For the **network-based similarity**, we calculate the shortest path length (SPL) similarity between aggregated phenotype terms *P* and candidate gene *g* as follows:

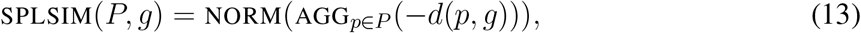

where *P* is the patient’s phenotype terms and *g* is a candidate gene, AGG is some aggregation function (*e*.*g*. mean),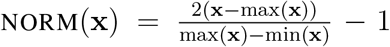 is a normalization function to scale the values in the range [*−*1, 1], and *d*(*p, g*) is the minimum number of hops between *p* and *g* in the KG. Whereas EMBSIM captures global network topology, SPLSIM captures complementary local network information via average shortest path lengths between the patient’s phenotypes and each candidate gene.

The final score between a patient’s phenotype terms *P* and candidate gene *g* (or **overall similarity**) is defined as:

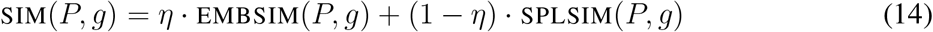

where *η* is a hyperparameter ranging from [0, 1] that represents the amount of weight to place on EMBSIM versus SPLSIM in the final gene prioritization scoring. SIM values range between [*−*1, 1].

For our **objective function**, we leverage a multi-similarity loss to encourage patient phenotype embeddings to be near their causal gene embedding and far away from the incorrect candidate gene embeddings. The multi-similarity loss is defined as follows [117]:

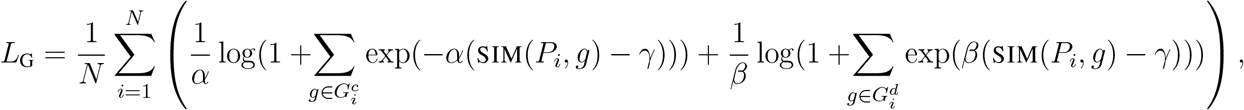

where *N* is the number of patients, *α, β*, and *γ* are hyperparameters, and SIM(*P*_*i*_, *g*) denotes the similarity between the aggregated phenotype embedding for patient *i* and the gene embedding of either the patient’s causal gene (*g ∈ G*^*c*^) or distractor gene (*g ∈ G*^*d*^). The optimized embedding space encodes patient information such that the similarity between a patient’s phenotype terms and candidate genes (*i*.*e*., how likely it is that a given gene explains the patient’s symptoms) is inversely proportional to the distance between the patient embedding and the embedding of the candidate gene.

### 12 SHEPHERD: Finding Similar Patients

SHEPHERD can find similar patients from a cohort of rare disease patients. This is important for identifying molecular diagnoses and validating already prioritized candidate genes. To match rare disease patients, SHEPHERD first learns a task-specific similarity function that captures the similarity between two patients. This training process produces an embedding space in which the similarity between two patients is inversely proportional to the distance between the two patient embeddings. The embedding space can be used at inference time to answer “patients-like-me” queries. We define the similarity between two patients *i* and *j* as the L2 distance between their aggregated phenotype embeddings 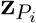 and 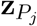:

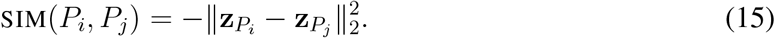

Importantly, when calculating patient similarity, we do not include any genotype information for the patients. This makes the model applicable in settings where the patient’s genome has not been sequenced or when the analysis results are still pending.

Regarding the **objective function**, SHEPHERD is trained to capture patient similarity using the neighborhood component analysis (NCA) loss:

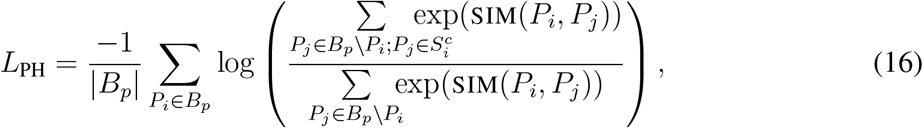

where *B*_*p*_ is a batch of patients sampled from the training set and *S*^*c*^ is the set of patients with the same causal gene as patient *P*_*i*_. Optimizing the NCA loss [118] minimizes the distances between patient embeddings with the same causal gene and maximizes the distances between patient embeddings with different causal genes.

### 13 SHEPHERD: Estimating Patient-Disease Similarity

Finally, SHEPHERD can characterize a clinical presentation based on existing knowledge about other rare and common diseases. We analogously perform novel disease characterization by learning an embedding space such that the similarity between a patient and a disease (*i*.*e*. how likely it is that a patient has that disease) is inversely proportional to the distance between the patient embedding and the disease embedding. We define the similarity between a patient’s phenotype terms *P* and disease *d* as the L2 distance between the aggregated phenotype embedding and the disease embedding:

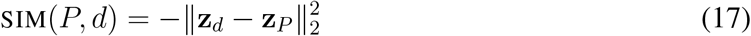

Regarding the **objective function** to optimize patient phenotype embeddings to be near their correct disease(s)’, we leverage a multi-modal version of the NCA loss, defined as:

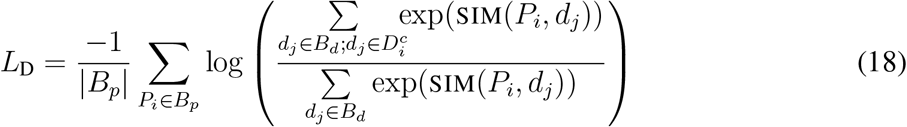

where *B*_*p*_ and *B*_*d*_ are batches of patients and candidate diseases, respectively, that are sampled from the training set, *P*_*i*_ corresponds to the phenotype term set for patient *i*, and 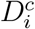 is the set of correct diseases for patient *i*. While 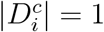 for most patients in our cohorts, several patients with multiple diseases exist.

### 14 Overall Objective Function

We train SHEPHERD in two stages. In the first stage, we pretrain the model to learn to capture the relationships between biomedical entities in the rare disease knowledge graph via self-supervised link prediction (*L*_LP_) only (Section 9). In the second stage, we finetune the pretrained model by simultaneously predicting relationships in the KG (*L*_LP_) and performing patient-centric rare disease tasks (*L*_DX_ *∈ {L*_G_, *L*_PH_, *L*_D_*}*) (Section 11-13). In other words, the pretrained model is jointly finetuned to achieve two distinct objectives: (1) to capture the relationships in the underlying knowledge graph and (2) to match a patient’s presenting symptoms with the patient’s causal gene(s), disease(s), or other similar patients. We model these objectives with two separate loss functions, link prediction loss *L*_LP_, which continues updating node embeddings, and diagnosis loss *L*_DX_ *∈ {L*_G_, *L*_D_, *L*_PH_*}*, which aligns patient phenotype terms to genes, diseases, or other patient phenotypes, respectively. The overall loss during the finetuning stage is as follows:

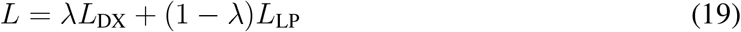

where *λ* is a hyperparameter controlling the weight of each loss. Whereas during pretraining, we train the model to capture generalizable biomedical knowledge by performing link prediction for all relation types, during fine-tuning, we focus on predicting gene, phenotype, and disease relations, which are most important for rare diseases. Training the model to perform link prediction enables the model to generalize to phenotypes and genes unseen in the training data.

### 15 Negative Sampling

To learn a meaningful representation space, we need negative examples (*i*.*e*. edges that do not exist in the KG, or candidate genes, diseases, or other patients that are not associated with a given patient). The following outlines the negative sampling strategies used for pretraining and each of the three rare disease diagnosis tasks.

For **link prediction**, we construct negative examples of triplets (*u, r, v*^*−*^) that do not exist in the KG by perturbing the target *nodes* while preserving the *types* of the source and target nodes and edge relation. For example, given a positive example of a triplet where the node and relation types are (*protein, has phenotype, phenotype*), a negative example is obtained by shuffling all phenotype *nodes* in the batch, thereby maintaining the node and relation *types* of the positive example.

For **causal gene discovery**, negative examples are constructed by taking the union of the candidate genes for all patients in a given batch. As noted in Section 3 to 6, each patient has a list of candidate genes that have been shortlisted as the most probable genes to cause the patient’s symptoms, and identifying the true causal gene(s) among them is especially challenging. We ensure that these “hard” candidate genes are included in the candidate list for each patient during training, as using such “hard” examples tends to improve the efficiency of training [119]. Furthermore, to maximize the number and frequency of candidate genes seen during train time, we up-sample a subset of candidate genes that are under-represented across all patients. Concretely, we count the frequencies of candidate genes in the prior and current batches, select the *k* most infrequently seen candidate genes (*i*.*e*. the *k* rarest candidate genes) in training batches, and add them to each patient’s candidate gene list. Note that we only prioritize the “hard” candidate genes for each patient at inference time without any up-sampling.

For **novel disease characterization**, negative examples include all diseases that do not explain the patient’s symptoms. First, we randomly sample 1,000 diseases from all diseases in the KG to serve as negative examples for each batch. Then, we calculate a patient’s similarity to all disease nodes in the KG at inference time. Due to the hierarchical structure of the KG, this approach may occasionally select parent or child diseases as negatives, introducing potential false negatives. However, the likelihood of sampling direct parent-child disease pairs as negatives is relatively low.

For **“patients-like-me” identification**, negative examples are simply all of the patients in the batch who do not have the same causal gene as the query patient. We construct batches to ensure at least two positive examples (*i*.*e*., patients with the same causal gene) for each patient in the batch. All remaining patients serve as negative examples. At inference time, we calculate a patient’s similarity to all patients in the cohort.

### 16 Disease-Split Training on Simulated and Publicly Available Patients

We train our model primarily on the simulated patient dataset. Training on simulated data offers several benefits: the simulated cohort is larger and more diverse than any real-world patient dataset, the trained models can be released without the risk of exposing any patient information, and the models can be evaluated on an independent real-world cohort to test how well a model can generalize to patients unseen during training. Further, and most importantly, we achieve generalizability to real-world cohorts by splitting patients into train and validation sets according to disease. Concretely, we split the list of diseases represented by the simulated patient cohort into train and validation. Then, we assign patients to train or validation sets such that patients with the same disease are either entirely in the training or fully in the validation set. As a result, the model is optimized so that its parameters are broadly transferable to patients with different diseases. The resulting train and validation cohorts contain 36,224 and 6,400 patients, respectively.

For the causal gene discovery task, we perform additional training on patients from the My-Gene2 and DDD cohorts. These additional cohorts constitute 3.6% of the training data. Unlike the UDN cohort, the MyGene2 and DDD cohorts do not have candidate genes for each patient. Therefore, we construct candidate gene lists by sampling 20 genes that are neighbors of each patient’s causal gene or phenotypes in the rare disease knowledge graph.

### 17 Additional Training Details

We provide details about node pretraining data splits, patient-driven sampling, and normalization.

Regarding **node pretraining data split**, edges in the knowledge graph are randomly split into train (80%), validation (10%), and test (10%) sets. Note that the forward and reverse edges of the same pair of nodes are maintained in the same data split to prevent data leakage.

For **patient-driven sampling**, we design a new approach for batch sampling that enables the model to perform patient gene prioritization while maintaining the topology of the KG. We first sample *m* patients and add their associated phenotypes and genes to the batch. Then, we add *n* nodes randomly sampled from the genes, phenotypes, and disease nodes in the KG. This allows for inductive generalization by maintaining the topology of nodes not found in the training data.

To help optimize model performance and convergence, we apply two **normalization** strategies to SHEPHERD. Specifically, we use LayerNorm [120] immediately after each convolutional layer and BatchNorm [121] following a nonlinear activation layer (here, leaky ReLU).

### 18 Hyperparameter tuning

We leverage Weights and Biases [122] to select optimal hyperparameters via a random search over the hyperparameter space. We first choose pretraining hyperparameters to optimize the micro F1 score on the pretraining validation set. The pretraining validation set consists of a set of edges that exist in the knowledge graph and a set of edges generated via negative sampling that do not exist (Section 15). Hyperparameters are selected via random search from the following values: learning rate *∈* [0.0001, 0.0005, 0.001, 0.005], weight decay *∈* [0, 0.005, 0.0005], dropout *∈* [0.2, 0.3, 0.4, 0.5, 0.6, 0.7, 0.8], and the number of GAT attention heads *∈* [2, 4]. We also perform a search over the dimension of the network layers: input size *∈* [2048, 4096], hidden size *∈* [256, 512, 1024], and output size *∈* [64, 128]. We then freeze the pretraining hyperparameters and perform a hyperparameter search independently for each rare disease task. We select task-specific hyperparameters to optimize the mean reciprocal rank of the correct genes, diseases, or patients on the disease-split simulated validation set. Importantly, the validation set containing simulated patients is entirely independent of the evaluation dataset, which includes patients from the Undiagnosed Diseases Network. We consider the following hyperparameters: learning rate *∈* [0.00001, 0.00005, 0.0001, 0.0005, 0.001], *λ ∈* [0.1, 0.9], *η ∈* [0.1, 0.9], *k* most infrequently seen genes *∈* [64, 128, 192], number of transformer layers for the phenotype encoder *∈* [0, 3, 6], number of heads in the transformer layers *∈* [4, 8, 16], contribution of the augmented gene embedding *ω ∈* [0.1, 0.9], number of *K* most similar genes to augment the gene embedding *∈* [1, 2, 3, 4, 5], and number of nodes *n* to sample per batch in *∈* [100, 200, 300, 400]. The code for hyperparameter selection and the optimal hyperparameters can be found at https://github.com/mims-harvard/SHEPHERD.

### 19 Implementation

We implement SHEPHERD using Pytorch (Version 1.8.0) [123], Pytorch Lightning (Version 1.4.5) [124], and Pytorch Geometric (Version 1.7.2) [125]. We leverage the Weights and Biases [122] platform for hyperparameter tuning and model training visualization, and we create interactive demos of the model using Gradio [126]. Models are trained on a single NVIDIA Quadro RTX8000 GPU. Training consists of approximately 15 hours of pretraining and 12 hours of fine-tuning, with early stopping based on validation performance; these times represent upper bounds, as the best-performing model typically emerges earlier in training. At inference time, SHEPHERD can run on a CPU, allowing it to efficiently rank any number of candidate genes without GPU memory constraints. SHEPHERD processes each patient in 3.46 seconds on average using the EXPERT-CURATED gene list (108 GB memory) and 3.97 seconds on average using the VARIANT-FILTERED gene list (109 GB memory). This enables efficient deployment in real-world clinical settings, including those with limited GPU resources.

### 20 Performance Stratified by Patient and Site Characteristics

We evaluate the trained model on the cohort of real-world UDN patients who have received a molecular diagnosis (Section 3). We measure the mean reciprocal rank of all of the patients’ causal genes and calculate the percentage of causal genes that appear in the top *k* ranked genes for *k ∈ {*1, 3, 5*}* for the EXPERT-CURATED candidate gene lists and *k ∈ {*1, 5, 10, 25, 50*}* for the longer VARIANT-FILTERED candidate gene lists. We analyze the performance across each of the UDN clinical sites, disease categories, and evaluation years. We also assess the correlation between model performance and the number of patient phenotype terms, the distance between the causal gene and phenotype terms in the KG, and the prevalence of the genetic conditions in the population. We leverage the number of ClinVar submissions for the causal gene as a proxy for prevalence.

### 21 Comparison to Alternative Approaches

We compare SHEPHERD to several diverse approaches for causal gene discovery. The first category includes network science or machine learning methods that enable us to assess the utility of SHEPHERD’s graph neural network approach and the use of simulated patients: (1) mean shortest graph distance, a network science approach that prioritizes genes according to their average shortest path in the KG to all of a patient’s phenotype terms; (2) supervised graph embedding, a logistic regression approach that frames prioritization as a binary prediction task for each candidate gene and represents each patient-gene option as the concatenation of the candidate gene’s pretrained node embedding and the patient’s averaged phenotype node embeddings; (3) supervised PCA embedding, a logistic regression approach similar to (2) that, instead of the KG node embeddings, utilizes a PCA-transformed shortest path length matrix from gene nodes to gene, phenotype, and disease nodes; and (4) large language models, namely the open-source LlaMa 3.1 8B and 70B parameter models [55].

We also compare SHEPHERD to six phenotype-based methods designed for causal gene discovery. Three are information-theoretic approaches that compare patient phenotype terms to either the phenotype terms associated with each candidate gene (ERIC, which is implemented in XRare [25]) or to the phenotype terms of all the diseases related to a candidate gene (Phrank [16] and PhenIX, which is implemented in Exomiser [24]). In the latter two approaches, each candidate gene is prioritized according to the highest similarity score across all associated diseases. LIRICAL is an approach that uses a likelihood ratio framework to estimate the extent to which patients’ phenotypes and genotypes are consistent across all known diseases [21]. CADA is a shallow network embedding-based approach that frames the task as link prediction between phenotype and gene nodes [19]. HiPhive is an approach implemented in Exomiser that leverages ontologies from humans and model organisms to assess phenotypic similarity. When phenotypic data is unavailable for a candidate, HiPhive employs a random walk approach on a protein-protein interaction network to establish connections between the candidate and other genes with similar phenotypes [24]. These baselines constitute a diverse set of methodological approaches for rare disease diagnosis. LIRICAL, XRare, and Exomiser are the strongest performing approaches in a prior comparison of gene prioritization tools [127], and Exomiser is currently in use at multiple UDN sites [50]. Finally, we compare to a non-guided baseline, Random, which represents performance without prioritizing candidate genes.

When possible, we leverage publicly available code and our KG to implement each causal gene discovery benchmark to elucidate whether performance differences were due to the algorithmic approach rather than a different or more up-to-date underlying knowledge base. In instances where we are unable to leverage our KG (*i*.*e*., for LIRICAL, HiPhive, and PhenIX), we leverage input data that includes only the gene-phenotype-disease associations that were present at the time when our rare disease KG was constructed to enable a fairer comparison. We use code from https://bitbucket.org/bejerano/phrank and https://github.com/Chengyao-Peng/CADA to run Phrank and CADA, respectively, using our rare disease KG. We run Exomiser v13.0.1 (released Nov 23, 2021) using their time-stamped input data from https://github.com/exomiser/Exomiser/tree/13.0.1 to ensure that we only utilize the associations known at the time of our KG construction. We run Exomiser with two different phenotype similarity options, PhenIX and HiPhive, and we leverage the `GENE PHENO SCORE` for prioritization. We re-implement the ERIC phenotype similarity score used in XRare; direct gene-phenotype edges and indirect gene-disease-phenotype paths from our KG are used to construct the phenotype terms associated with each candidate gene. We run LIRICAL with the “orphanet” data flag using code from https://github.com/TheJacksonLaboratory/ LIRICAL by supplying both a VCF file and positive and negative (*i*.*e*., that the patient did not exhibit) phenotype terms. Notably, all baselines except for LIRICAL leverage Exomiser to generate the VARIANT-FILTERED candidate genes for each patient before phenotype-based prioritization. In contrast, LIRICAL performs its variant-based filtering using the provided VCF files.

We run LlaMa 3.1 8B and 70B models using the Ollama Python library (https://github.com/ollama/ollama). We construct a prompt that instructs the model to use its “knowledge of genetics, known disease-gene associations, and variant interpretation” to generate a ranked list of all of the candidate genes based on their likelihood of causing the patient’s symptoms. The complete prompt is detailed in Supplementary Note 1. HPO terms are included in the prompt as textual descriptions of phenotypes, and candidate genes are provided as Ensembl IDs or Gene Symbols. We evaluate model performance under two conditions: when supplied with both phenotype terms and candidate genes, and when supplied only phenotype terms. If a candidate gene list is too long, the model will frequently refuse to rank the list. To address this, we split long lists containing at least 500 genes into two smaller subsets, rank each separately, and then use a prompt to merge the ranked lists (see Supplementary Note 1). If the model refuses to rank the shorter lists, we default to ranking based solely on phenotype descriptions. All experiments use the default hyperparameters provided by the Ollama OpenAI-compatible API.

We also compare SHEPHERD to approaches that can be used to identify similar patients. The information theoretic approach Phrank can be leveraged to calculate the semantic similarity between two sets of patient phenotype terms based on the information content of their shared phenotype ancestors in the Human Phenotype Ontology [16]. SET BASED calculates distance between two sets of phenotype terms *P* and *P* using Jaccard distance, defined as 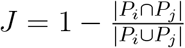.

We further compare SHEPHERD to a network-based phenotype search approach for novel disease characterization. For each patient phenotype, we identify its associated diseases based on our KG (*i*.*e*., direct disease neighbors of each phenotype node) and retrieve the disease category of the diseases. The percent similarity of the patient’s disease presentation to each disease category is computed as the percentage of the associated diseases in that category. These similarities become the KG-derived interpretable name for the patient’s novel disease presentation.

### 22 Assessing Statistical Significance

We perform a one-sided Wilcoxon signed-rank test to assess whether there is a significant difference in causal gene discovery performance between SHEPHERD and baseline methods. After confirming that the data is not normally distributed, we evaluate whether there is a statistically significant difference in SHEPHERD’s performance across clinical sites, evaluation years, and primary presenting symptoms using a Kruskal-Wallis H-test. In the knowledge graph, we calculate the Spearman correlation coefficient to measure the correlation between causal gene rank and the distance between a patient’s phenotype terms and causal gene. To assess whether patients cluster by disease category, we perform K-means clustering with *k* set to the number of disease categories, and we evaluate the clusters according to an adjusted mutual information score from `scikit-learn`, which is designed to consider clusters of different sizes. We assess the significance of the resulting clustering via a permutation test with 100 random permutations of the true cluster labels. We perform a Mann-Whitney test to measure the difference in distances in the embedding space for patients with the same versus different disease categories. Finally, we perform the two-sample Kolmogorov-Smirnov test to assess whether the distribution of embedding distances for patients with the same disease is identical to that for patients with different diseases.

### 23 Visualization of Learned Embeddings

We visualize embeddings learned via SHEPHERD in a Uniform Manifold Approximation and Projection (UMAP) plot [128]. We use the `umap-learn` Python package [129] and perform a grid search over the `n neighbors`, `min dist`, and `spread` UMAP parameters. We select parameters that maintain global structure.

### 24 Visualization of Patient Neighborhoods in the Knowledge Graph

To visualize the local neighborhood of patients’ disease, phenotype, and gene nodes (Figure 4), we calculate the shortest paths between patient-relevant nodes and extract all nodes in those shortest paths. We visualize the resulting patient neighborhoods using Gephi 0.9.4 [130]. We apply Fruchterman Reingold, Noverlap, and Label Adjust layouts and manual adjustment to organize the nodes so they do not overlap.

## Data availability

All data used in the paper, including the rare disease knowledge graph, simulated, MyGene2, and DDD patient cohorts, and the final and intermediate results of the analyses are shared with the research community at https://zitniklab.hms.harvard.edu/projects/SHEPHERD. The patient dataset derived from the Deciphering Developmental Disorders Study (DDD) is created using aggregated gene and phenotypic information from patients in the Deciphering Developmental Disorders Study (https://www.deciphergenomics.org/ddd/ddgenes), an initiative of nearly 14,000 pediatric patients with severe undiagnosed developmental disorders from the United Kingdom and Ireland. While the UDN dataset cannot be released due to privacy concerns, anonymized UDN data has been deposited in dbGaP (accession phs001232) and PhenomeCentral. Phenotypes and causal variants and genes related to UDN diagnoses are also shared publicly in ClinVar at https://www.ncbi.nlm.nih.gov/clinvar/submitters/505999. The UDN study is approved by the NIH IRB Protocol 15HG0130. All patients accepted to the UDN provide written informed consent to share their data across the UDN.

## Code availability

Python implementation of the methodology developed and used in the study is available via the project website at https://zitniklab.hms.harvard.edu/projects/SHEPHERD. The code to reproduce results, documentation, and examples are at https://github.com/mims-harvard/SHEPHERD. We provide an interactive tool to explore SHEPHERD’s outputs at https://huggingface.co/spaces/emilyalsentzer/SHEPHERD.

## Acknowledgements

We thank Kimberly LeBlanc for reviewing our work to ensure we follow UDN data privacy protocols. E.A. is supported by a Microsoft Research PhD Fellowship. M.M.L. is supported by T32HG002295 from the National Human Genome Research Institute and a National Science Foundation Graduate Research Fellowship. M.Z. gratefully acknowledges the support by NSF under Nos. IIS-2030459 and IIS-2033384, US Air Force Contract No. FA8702-15-D-0001, and awards from Harvard Data Science Initiative, Amazon Research, Bayer Early Excellence in Science, AstraZeneca Research, and Roche Alliance with Distinguished Scientists. UDN research reported in this manuscript was supported by the NIH Common Fund, through the Office of Strategic Coordination/Office of the NIH Director under the following award numbers: U01HG007709, U01HG010219, U01HG010230, U01HG010217, U01HG010233, U01HG010215, U01HG007672, U01HG007690, U01HG007708, U01HG007703, U01HG007674, U01HG007530, U01HG007942, U01HG007943, U01TR001395, U01TR002471, U54NS108251, and U54NS093793. This study also makes use of data generated by the DECIPHER community. A full list of centers who contributed to the generation of the data is available from https://deciphergenomics.org/about/stats and via email from. DECIPHER is hosted by EMBL-EBI and funding for the DECIPHER project was provided by the Wellcome Trust [grant number WT223718/Z/21/Z]. The content is solely the responsibility of the authors and does not necessarily represent the official views of the National Institutes of Health and other funders.

## Authors contributions

E.A., M.M.L., and S.K. retrieved and processed the UDN, MyGene2, and simulated patient data. E.A. and M.M.L. developed, implemented, and benchmarked SHEPHERD and performed detailed analyses of SHEPHERD’s algorithm. E.A., M.M.L., I.K., and M.Z. designed the study. E.A., M.M.L., S.K., A.N., I.K., and M.Z. contributed to writing the manuscript.

