## Supplementary Information for "Few shot learning for phenotype-driven diagnosis of patients with rare genetic diseases"

### Supplementary Figures and Tables

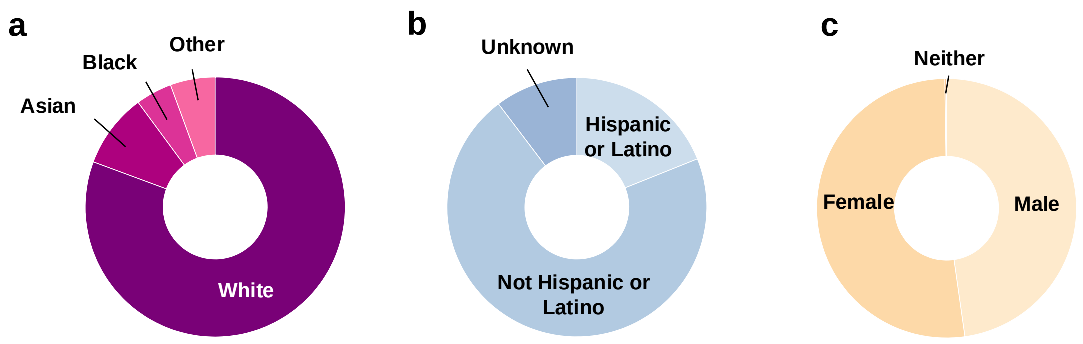


**Supplementary Figure 1: UDN cohort demographics.** Number of patients with each (a) race, (b) ethnicity, and (c) sex among UDN patients.

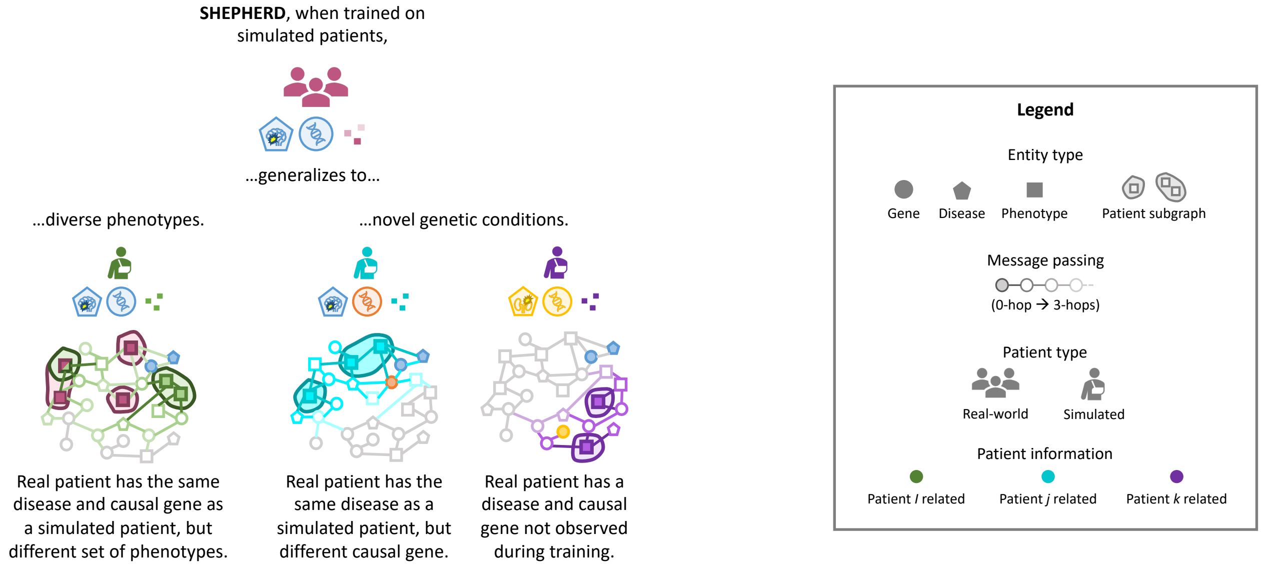


**Supplementary Figure 2: SHEPHERD can generalize to heterogeneous phenotypic presentations and novel genetic conditions.** There are few patients with each rare disease, and patients with the same disease can have variable clinical presentations. SHEPHERD is trained on simulated rare disease patients and can generalize to real-world patients with unique, unseen phenotypes (left), with novel disease-causing genes (center), and with entirely novel diseases (right).

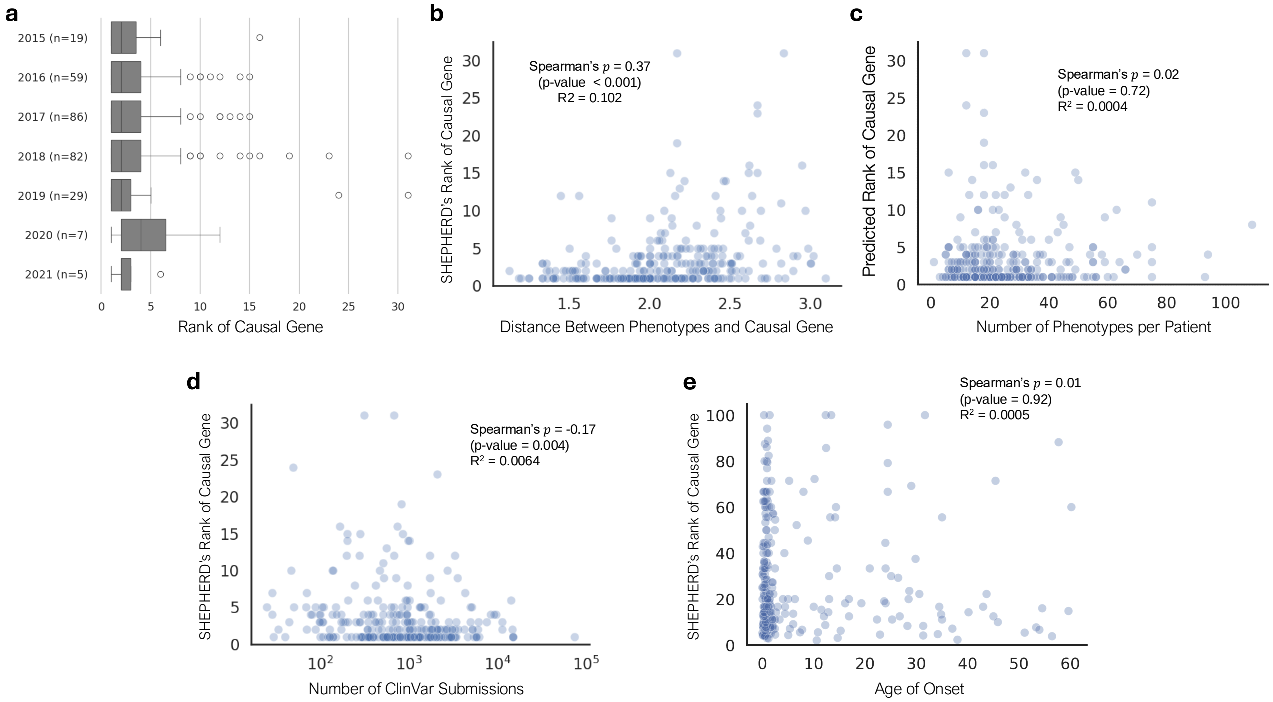


**Supplementary Figure 3: Generalizability of causal gene discovery performance on EXPERT-CURATED candidate lists.** **(a)** Performance of SHEPHERD in ranking causal genes stratified by evaluation year on the EXPERT-CURATED gene list **(b)** Correlation between model performance (i.e., the rank of a disease-driving gene) and the average distance between a patient's phenotypes and causal genes in the knowledge graph. **(c)** Correlation between model performance and the number of phenotype terms describing each patient's clinical presentation. **(d)** Correlation between model performance and prevalence of the rare genetic disorders. The number of submissions to the database ClinVar for the causal gene is used as a surrogate for the prevalence of the rare disorders. The x-axis is number of submissions in log-scale. **(e)** Correlation between model performance and the age of onset.

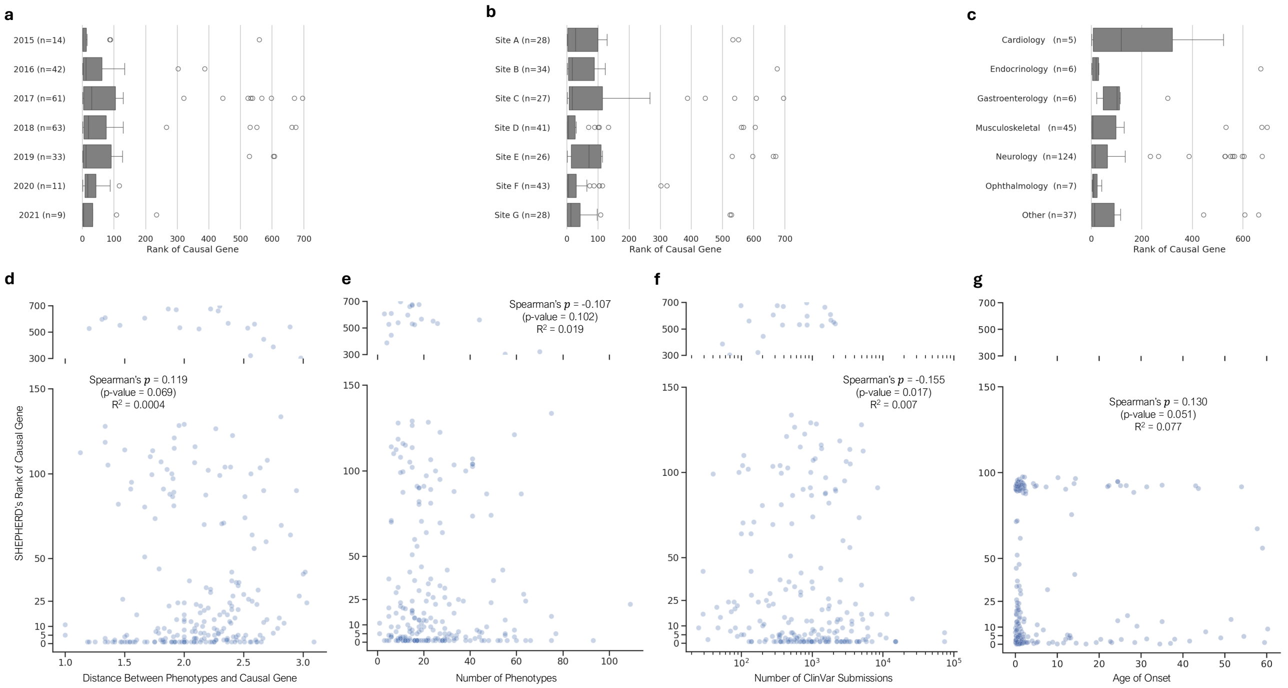


**Supplementary Figure 4: Generalizability of causal gene discovery performance on VARIANT-FILTERED candidate lists.** **(a-c)** Performance of SHEPHERD in ranking causal genes stratified by **(a)** evaluation year, **(b)** clinical site, and **(c)** primary presenting symptom on the VARIANT-FILTERED gene list **(d)** Correlation between model performance (i.e., the rank of a disease-driving gene) and the average distance between a patient's phenotypes and causal genes in the knowledge graph. **(e)** Correlation between model performance and the number of phenotype terms describing each patient's clinical presentation. **(f)** Correlation between model performance and prevalence of the rare genetic disorders. **(g)** Correlation between model performance and the age of onset. The number of submissions to the database ClinVar for the causal gene is used as a surrogate for the prevalence of the rare disorders. The x-axis is number of submissions in log-scale.

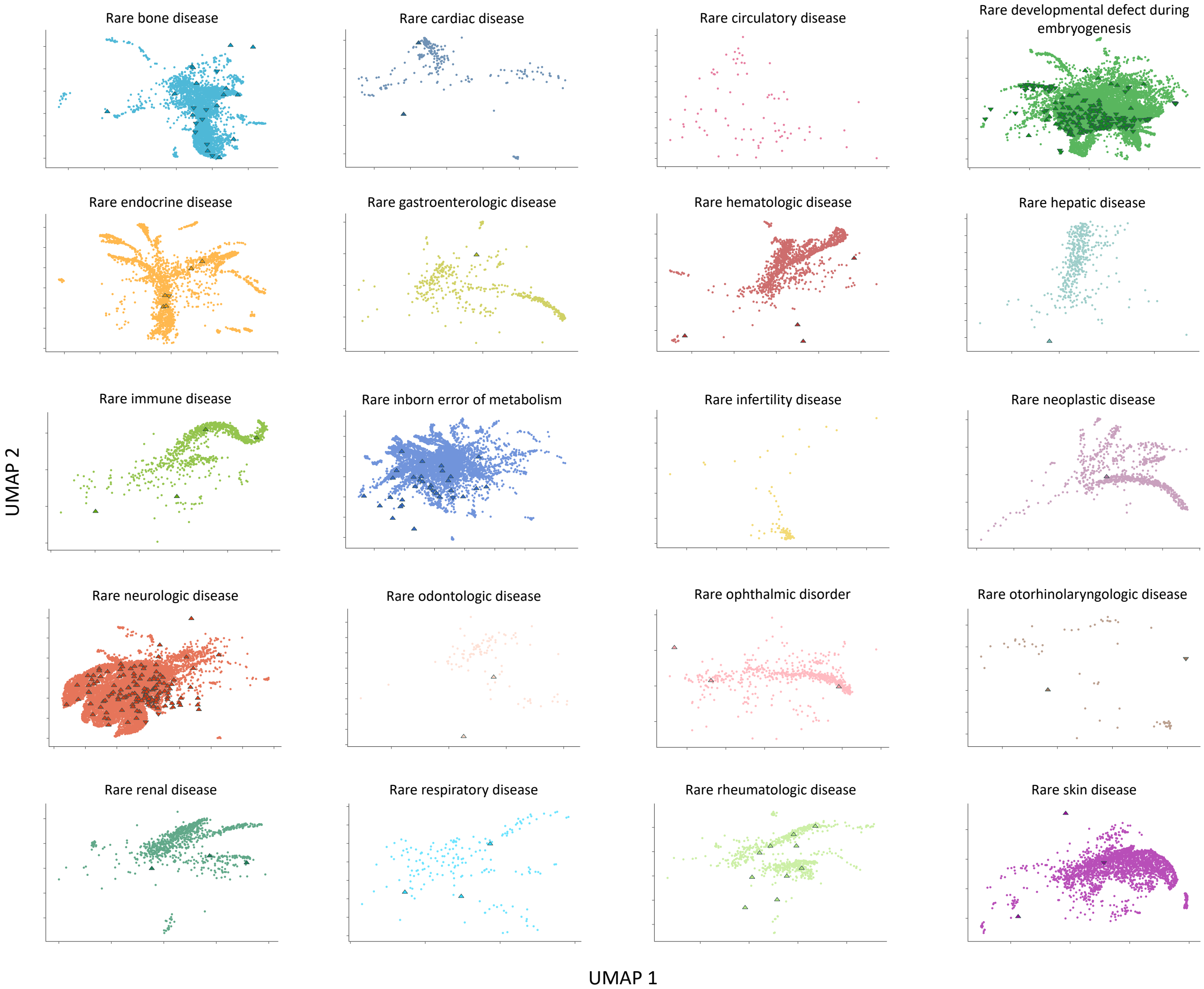


**Supplementary Figure 5: Visualization of rare disease patients by disease category.** Two-dimensional UMAP plot of SHEPHERD's embedding space of all simulated patients (circles) and two real-world cohorts of UDN patients (up-facing triangles) and MyGene2 patients (down-facing triangles) grouped by the Orphanet disease category of medical diagnosis. Simulated, MyGene2 and UDN patients embed nearby other patients whose diagnoses belong to the same disease category.

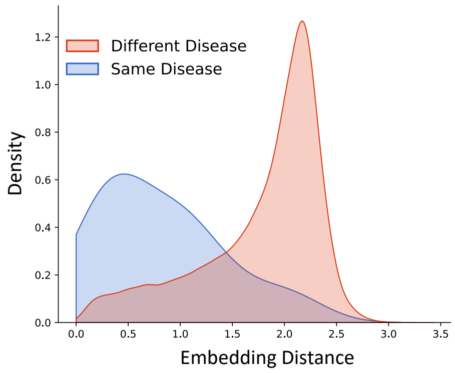


**Supplementary Figure 6: SHEPHERD performs patients-like-me identification.** Distribution of SHEPHERD embedding distance between UDN and MyGene2 patients with the same vs. different diseases.

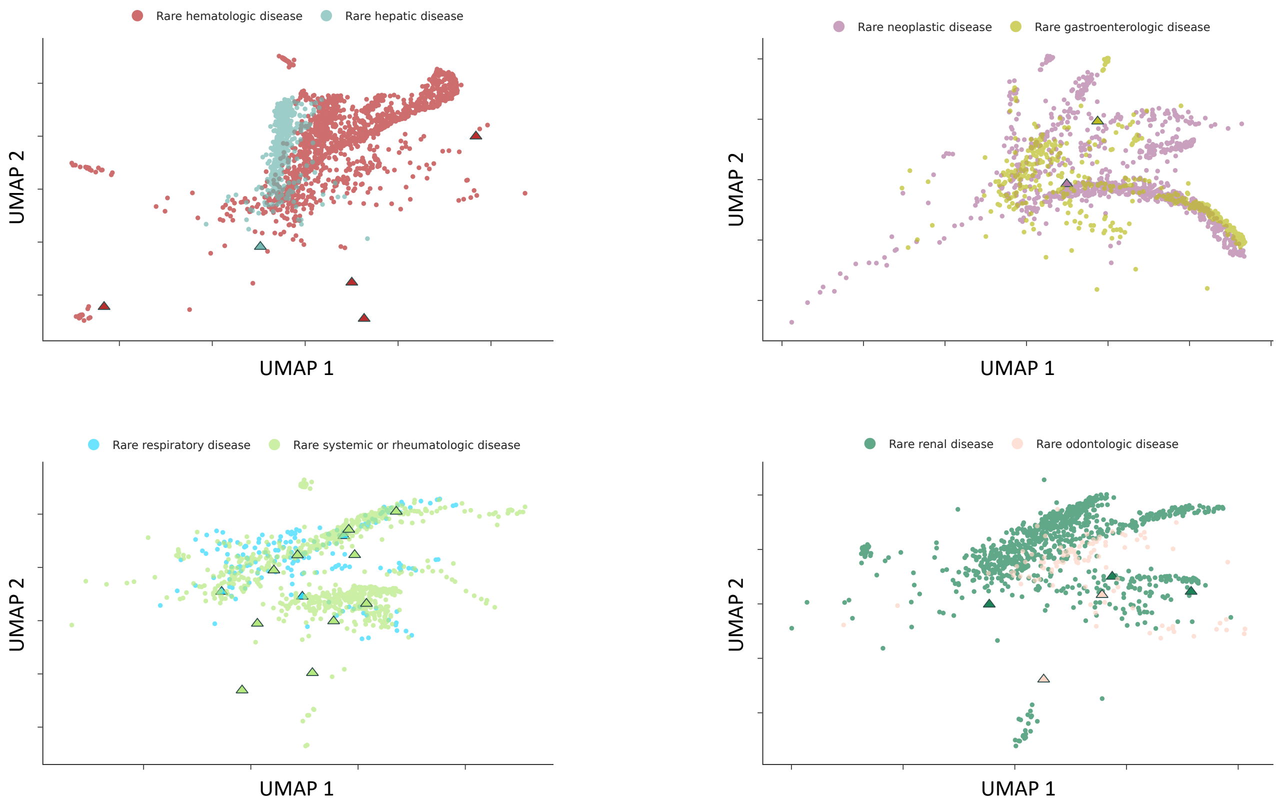


**Supplementary Figure 7: Visualization of the relationship between disease categories.** Two-dimensional UMAP plot of SHEPHERD's embedding space for the most similar pairs of disease categories. Circles correspond to simulated patients, up-facing triangles to UDN patients, and down-facing triangles to MyGene2 patients.

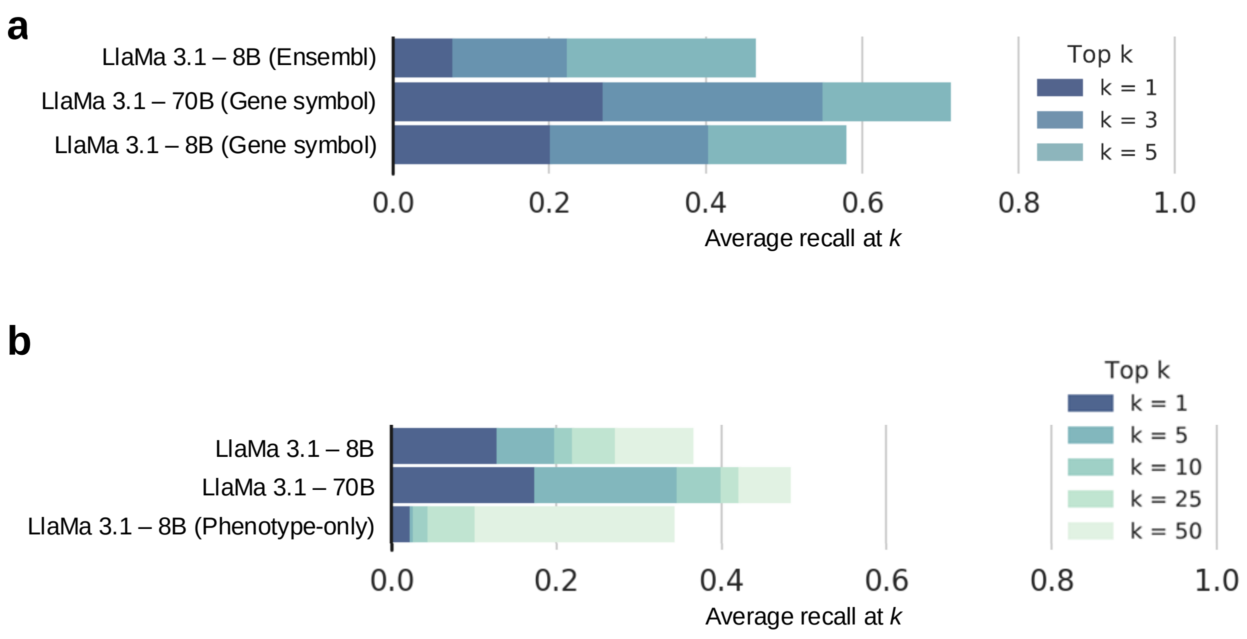


**Supplementary Figure 8: Evaluation of LlaMa 3.1 models.** Performance of different LlaMa 3.1 8B and 70B models on **(a)** the EXPERT-CURATED gene lists, where genes are kept as Ensembl IDs or mapped to gene symbols, and **(b)** the VARIANT-FILTERED gene lists or all genes (i.e., phenotype-only).

**Supplementary Table 1:** Non-overlapping disorders between all phenotyped patients in the Undiagnosed Diseases Network (UDN) and simulated (SIM) patient cohorts. The names of the 5 most frequently observed diseases that are not in the other patient cohort are shown. The full list of syndromes found across all cohorts can be found in the Harvard Dataverse Repository at the following link: https://dataverse.harvard.edu/file.xhtml?fileId=10214709&version=3.0.

| Rank | Diseases in UDN but not in SIM | Diseases in SIM but not in UDN |
|---|---|---|
| 1 | neurodevelopmental disorder with regression, abnormal movements, loss of speech, and seizures | multiple intestinal atresia |
| 2 | TBCK-related intellectual disability syndrome | progeroid syndrome, Petty type |
| 3 | dystonia 28, childhood-onset | myofibrillar myopathy 3 |
| 4 | Rett syndrome, congenital variant | GM3 synthase deficiency |
| 5 | Bethlem myopathy 1 | otospondylomegaepiphyseal dysplasia, autosomal dominant |

### Supplementary Notes

**Supplementary Note 1: Causal gene discovery prompts for LlaMa 3.1 models.**

We use three types of prompts in our experiments with the LlaMa 3.1 models.

**(a) Prompt used when the patient's phenotypes and candidate genes are both provided as input.**

You are an expert in rare disease diagnosis. I will provide a list of Human Phenotype Ontology (HPO) terms describing a patient's symptoms, along with a list of candidate genes. Using your knowledge of genetics, known disease-gene associations, and variant interpretation, generate a ranked list of all of the candidate genes based on their likelihood of causing the patient's symptoms. The output should be in JSON Lines (jsonl) format, with each line containing the gene name, rank, and a brief explanation of why the gene is relevant to the patient's HPO terms. Only output a valid jsonl file with no spaces between each json. Rank every candidate gene according to its association with the HPO terms, known gene-disease relationships, and functional impact. Make sure to rank all candidates.

Output (JSON Lines format):
{"gene_name": "Gene1", "rank": 1, "explanation": "Explanation for why Gene1 is relevant" }
{"gene_name": "Gene2", "rank": 2, "explanation": "Explanation for why Gene2 is relevant" }

**(b) Prompt used when only the patient's phenotypes are provided as input.**

You are an expert in rare disease diagnosis. I will provide a list of Human Phenotype Ontology (HPO) terms describing a patient's symptoms. Using your knowledge of genetics, known disease-gene associations, and variant interpretation, generate a ranked list of all of the candidate genes as Gene Symbols based on their likelihood of causing the patient's symptoms. The output should be in JSON Lines (jsonl) format, with each line containing the gene symbol, rank, and a brief explanation of why the gene is relevant to the patient's HPO terms. Only output a valid jsonl file with no spaces between each json. Rank every candidate gene according to its association with the HPO terms, known gene-disease relationships, and functional impact. Make sure to rank all candidates.

Output (JSON Lines format):
{ "gene_name": "Gene1", "rank": 1, "explanation": "Explanation for why Gene1 is relevant" }
{ "gene_name": "Gene2", "rank": 2, "explanation": "Explanation for why Gene2 is relevant" }

**(c) Prompt used to merge two previously ranked candidate gene lists.** *When patients have very long candidate gene lists, we split the lists in two, rank each smaller list, and use a language model* *to merge the two ranked lists into a final ranked list.*

You are an expert in rare disease diagnosis. I will provide Human Phenotype Ontology (HPO) terms describing a patient's symptoms, along with two lists of previously ranked candidate genes. Using your knowledge of genetics, known disease-gene associations, and variant interpretation, combine the two ranked lists of all of the candidate genes based on their likelihood of causing the patient's symptoms. The output should be in JSON Lines (jsonl) format, with each line containing the gene name, rank, and a brief explanation of why the gene is relevant to the patient's HPO terms. Only output a valid jsonl file with no spaces between each json. Rank every candidate gene according to its association with the HPO terms, known gene-disease relationships, and functional impact. Make sure to rank all candidates.

Output (JSON Lines format):
{"gene_name": "Gene1", "rank": 1, "explanation": "Explanation for why Gene1 is relevant" }
{"gene_name": "Gene2", "rank": 2, "explanation": "Explanation for why Gene2 is relevant" }

### Members of the Undiagnosed Diseases Network (Version: 3.31.25)

| Full Name | Affiliation |
| --- | --- |
| Aaron Quinlan | University of Utah |
| Abdul Elkadri | MCW-CW |
| Adeline Vanderver | CHOP |
| Adriana Rebelo | Miami |
| Alan H. Beggs | Harvard |
| Albert R. La Spada | UCI/CHOC |
| Alden Huang | UCLA |
| Alex Paul | WUSTL Clinical |
| Alexander Miller | Stanford |
| Ali Al-Beshri | UAB |
| Alistair Ward | University of Utah |
| Allen Bale | Yale |
| Allyn McConkie-Rosell | Duke |
| Alyssa A. Tran | BCM Clinical |
| Andrea Gropman | NIH UDP |
| Andres Vargas | UCLA |
| Andrew B. Crouse | UAB DMCC |
| Andrew Stergachis | PNW |
| Anna Hurst | UAB |
| Anna Raper | CHOP/UPenn |
| Arjun Tarakad | BCM Clinical |
| Ashley Andrews | University of Utah |
| Ashley McMinn | Vanderbilt |
| Ashok Balasubramanyam | BCM Clinical |
| Ayuko Iverson | Mount Sinai |
| Barbara N. Pusey Swerdzewski | NIH UDP |
| Beatriz Anguiano | Stanford |
| Ben Afzali | NIH UDP, NHGRI |
| Ben Solomon | NIH UDP, NHGRI |
| Beth A. Martin | Stanford |
| Bianca E. Russell | UCLA |
| Brandon M Wilk | UAB |
| Breanna Mitchell | Mayo Clinic |
| Brendan C. Lanpher | Mayo Clinic |
| Brendan H. Lee | BCM Clinical |
| Brent L. Fogel | UCLA |
| Brett Bordini | MCW-CW |

| | |
|---|---|
| Brett H. Graham | IU |
| Brian Corner | Vanderbilt |
| Brianna Tucker | Stanford |
| Bruce Gelb | Mount Sinai |
| Bruce Korf | UAB |
| Calum A. MacRae | Harvard |
| Camilo Toro | NIH UDP |
| Cara Skraban | CHOP |
| Carlos A. Bacino | BCM Clinical |
| Carol Oladele | Yale |
| Caroline Hendry | Yale |
| Carson A. Smith | Miami |
| Cecilia Esteves | Harvard DMCC |
| Changrui Xiao | UCI/CHOC |
| Charlotte Cunningham-Rundles | Mount Sinai |
| Chloe M. Reuter | Stanford |
| Christine M. Eng | BCM Sequencing |
| Chun-Hung Chan | Sanford |
| Colleen E. Wahl | NIH UDP |
| Corrine K. Welt | University of Utah |
| Cynthia J. Tifft | NIH UDP, NHGRI |
| Dana Kiley | WUSTL Clinical |
| Daniel J. Rader | CHOP/UPenn |
| Daniel Wegner | WUSTL Clinical |
| Danny Miller | PNW |
| Daryl A. Scott | BCM Clinical |
| Dave Viskochil | University of Utah |
| David A. Sweetser | Harvard |
| David R. Adams | NIH UDP, NHGRI |
| Deborah Barbouth | Miami |
| Deepak A. Rao | Harvard |
| Devin Oglesbee | Mayo Clinic |
| Devon Bonner | Stanford |
| Donald Basel | MCW-CW |
| Donna Novacic | NIH UDP |
| Dr. Francisco Bustos velasq | Sanford |
| Dustin Baldridge | WUSTL MOSC |
| Edward Behrens | CHOP |
| Edwin K. Silverman | Harvard |
| Elaine Seto | BCM Clinical |

| | |
|---|---|
| Elijah Kravets | Stanford |
| Elisabeth Rosenthal | PNW |
| Elizabeth A Worthey | UAB |
| Elizabeth A. Burke | NIH UDP, NHGRI |
| Elizabeth Blue | PNW |
| Elizabeth C. Chao | UCI/CHOC |
| Elizabeth L. Fieg | Harvard |
| Ellen F. Macnamara | NIH UDP |
| Elsa Balton | PNW |
| Emily Glanton | Harvard DMCC |
| Emily Shelkowitz | PNW |
| Emily Wang | Yale |
| Eric Allenspach | PNW |
| Eric Gayle | Mount Sinai |
| Eric Klee | Mayo Clinic |
| Eric Vilain | UCI/CHOC |
| Erin Conboy | IU |
| Erin E. Baldwin | University of Utah |
| Erin McRoy | WUSTL Clinical |
| Esteban C. Dell'Angelica | UCLA |
| Euan A. Ashley | Stanford DMCC |
| F. Sessions Cole | WUSTL DMCC |
| Filippo Pinto e Vairo | Mayo Clinic |
| Frances High | Harvard |
| Francesco Vetrini | IU |
| Francis Rossignol | NIH UDP, NHGRI |
| Fuki M. Hisama | PNW |
| Gabor Marth | University of Utah DMCC |
| Gail P. Jarvik | PNW |
| Gary D. Clark | BCM Clinical |
| George Carvalho | UCLA |
| Gerard T. Berry | Harvard |
| Ghayda Mirzaa | PNW |
| Giorgio Sirugo | CHOP/UPenn |
| Gonench Kilich | CHOP |
| Guney Bademci | Miami |
| Hector Rodrigo Mendez | Stanford |
| Heidi Wood | NIH UDP, NHGRI |
| Herman Taylor | Morehouse DMCC |
| Holly K. Tabor | Stanford |

| | |
|---|---|
| Hongzheng Dai | BCM Clinical |
| Hsiao-Tuan Chao | BCM Clinical |
| Hua Xu | Yale |
| Hugo J. Bellen | BCM MOSC |
| Hui Zhang | Yale |
| Ian Glass | PNW |
| Ian R. Lanza | Mayo Clinic |
| Ingrid A. Holm | Harvard |
| Isaac S. Kohane | Harvard DMCC |
| Isum Ward | Sanford |
| Ivan Chinn | BCM Clinical |
| J. Carl Pallais | Harvard |
| Jacinda B. Sampson | Stanford |
| James P. Orengo | BCM Clinical |
| James Verbsky | MCW-CW |
| Jared Sninsky | BCM Clinical |
| Jason Hom | Stanford |
| Jason Schend | Sanford |
| Jennefer N. Kohler | Stanford |
| Jennifer E. Posey | BCM Clinical |
| Jennifer Morgan | Sanford |
| Jennifer Schymick | Stanford |
| Jennifer Wambach | WUSTL Clinical |
| Jessica Douglas | Harvard |
| Jiayu Fu | NIH UDP, NHGRI |
| Jill A. Rosenfeld | BCM Clinical |
| Jimann Shin | WUSTL MOSC |
| Joan M. Stoler | Harvard |
| Joanna Jen | Mount Sinai |
| Joanna M. Gonzalez | Miami |
| John A. Phillips III | Vanderbilt |
| John Carey | University of Utah |
| John E. Gorzynski | Stanford |
| John J. Mulvihill | NIH UDP |
| Joie Davis | NIH UDP, NHGRI |
| Jonathan A. Bernstein | Stanford |
| Jordan Whitlock | UAB DMCC |
| Jose Abdenur | UCI/CHOC |
| Joseph Loscalzo | Harvard |
| Joy D. Cogan | Vanderbilt |

| | |
|---|---|
| Julian A. Martínez-Agosto | UCLA |
| Julie McCarrier | MCW-CW |
| Justin Alvey | University of Utah |
| Kahlen Darr | Mayo Clinic |
| Kaitlin Callaway | UAB |
| Kathleen A. Leppig | PNW |
| Kathleen Sullivan | CHOP |
| Kathy Sisco | WUSTL Clinical |
| Katrina Dipple | PNW |
| Kayla M. Treat | IU |
| Kelly Hassey | CHOP |
| Kelly Schoch | Duke |
| Kevin S. Smith | Stanford |
| Khurram Liaqat | IU |
| Kim Worley | BCM Clinical |
| Kimberly Ezell | Vanderbilt |
| Kimberly LeBlanc | Harvard DMCC |
| Kirsten Blanco | UCI/CHOC |
| Kumarie Latchman | Miami |
| Lance H. Rodan | Harvard |
| Laura Keehan | Stanford |
| Laura Pace | University of Utah |
| Laurel A. Cobban | Harvard |
| Lauren Blieden | BCM Clinical |
| Lauren C. Briere | Harvard |
| Lauren Jeffries | Yale |
| Laurens Wiel | Stanford |
| Layal F. Abi Farraj | UCLA |
| Leoyklang Petcharet | NIH UDP, NHGRI |
| LéShon Peart | Miami |
| Lili Mantcheva | IU |
| Lilianna Solnica-Krezel | WUSTL MOSC |
| Lindsay C. Burrage | BCM Clinical |
| Lindsay Mulvihill | Mayo Clinic |
| Lisa Schimmenti | Mayo Clinic |
| Lisa T. Emrick | BCM Clinical |
| Lorenzo Botto | University of Utah |
| Lorraine Potocki | BCM Clinical |
| Louise Bier | Mount Sinai |
| Lynette Rives | Vanderbilt |

| | |
|---|---|
| Lynne A. Wolfe | NIH UDP, NHGRI |
| Mafalda Barbosa | Mount Sinai |
| Maija-Rikka Steenari | UCI/CHOC |
| Manish J. Butte | UCLA |
| Manisha Balwani | Mount Sinai |
| Margaret Delgado | NIH UDP, NHGRI |
| María José Ortuño Romero | Yale |
| Maria T. Acosta | NIH UDP |
| Marie Morimoto | NIH UDP, NHGRI |
| Mariko Nakano-Okuno | UAB DMCC |
| Mariya Shadrina | Mount Sinai |
| Mark Gerstein | Yale |
| Mark Wener | PNW |
| Marla Sabaii | NIH UDP, NHGRI |
| Martha Horike-Pyne | PNW |
| Martin G. Martin | UCLA |
| Martin Rodriguez | UAB |
| Matt Velinder | University of Utah |
| Matthew Coggins | Harvard |
| Matthew Might | UAB DMCC |
| Matthew T. Wheeler | Stanford |
| MayChristine V. Malicdan | NIH UDP, NHGRI |
| Megan Bell | Sanford |
| Meghan C. Halley | Stanford |
| Melissa Walker | Harvard |
| Mia Levanto | Stanford |
| Michael Bamshad | PNW |
| Michael F. Wangler | BCM MOSC |
| Michael Muriello | MCW-CW |
| Michael Zimmermann | MCW-CW |
| Michele Spencer-Manzon | Yale |
| Miranda Leitheiser | Sanford |
| Mohamad Mikati | Duke |
| Mohamad Saifeddine | Sanford |
| Monika Weisz Hubshman | BCM Clinical |
| Monkol Lek | Yale |
| Monte Westerfield | UO MOSC |
| Mustafa Tekin | Miami |
| Nada Derar | Yale |
| Naghmeh Dorrani | UCLA |

| | |
|---|---|
| Neil H. Parker | UCLA |
| Neil Hanchard | NIH UDP, NHGRI |
| Nicholas Borja | Miami |
| Nicola Longo | University of Utah |
| Nicole M. Walley | Duke |
| Nitsuh K. Dargie | PNW |
| Odelya Kaufman | Yale |
| Oguz Kanca | BCM MOSC |
| Orpa Jean-Marie | NIH UDP, NHGRI |
| Page C. Goddard | Stanford |
| Paolo Moretti | University of Utah |
| Patricia A. Ward | BCM Sequencing |
| Patricia Dickson | WUSTL Clinical |
| Paul Berger | Sanford |
| Paul G. Fisher | Stanford |
| Pengfei Liu | BCM Sequencing |
| Peter Byers | PNW |
| Pinar Bayrak-Toydemir | University of Utah/ARUP |
| Precilla D'Souza | NIH UDP |
| Queenie Tan | Mayo Clinic |
| Rachel A. Ungar | Stanford |
| Rachel Evard | Mount Sinai |
| Rachel Li | Sanford |
| Rachel Mahoney | Harvard DMCC |
| Rakale C. Quarells | Morehouse DMCC |
| Ramakrishnan Rajagopalan | CHOP |
| Raquel L. Alvarez | Stanford |
| Rebecca C. Spillmann | Duke |
| Rebecca Ganetzky | CHOP |
| Rebecca Overbury | University of Utah |
| Rebekah Barrick | UCI/CHOC |
| Richard A. Lewis | BCM Clinical |
| Richard Chang | UCI/CHOC |
| Richard L. Maas | Harvard |
| Rizwan Hamid | Vanderbilt |
| Rong Mao | University of Utah/ARUP |
| Ronit Marom | BCM Clinical |
| Rosario I. Corona | UCLA |
| Runjun Kumar | PNW |
| Russell Butterfield | University of Utah |

| | |
|---|---|
| Sanaz Attaripour | UCI/CHOC |
| Sandesh Nagamani | BCM Clinical |
| Sara Emami | Stanford |
| Saskia Shuman | Mount Sinai |
| Seema R. Lalani | BCM Clinical |
| Serena Neumann | Vanderbilt |
| Seth Perlman | PNW |
| Shamika Ketkar | BCM Clinical |
| Shamil R. Sunyaev | Harvard DMCC |
| Shilpa N. Kobren | Harvard DMCC |
| Shinya Yamamoto | BCM MOSC |
| Shrikant Mane | Yale |
| Shruti Marwaha | Stanford |
| Sirisak Chanprasert | PNW |
| Stanley F. Nelson | UCLA |
| Stephan Zuchner | Miami |
| Stephanie Bivona | Miami |
| Stephanie M. Ware | IU |
| Stephen B Montgomery | Stanford |
| Stephen C. Pak | WUSTL MOSC |
| Steven Boyden | University of Utah |
| Suha Bachir | Stanford |
| Surendra Dasari | Mayo Clinic |
| Susan Korrick | Harvard |
| Susan Shin | Mount Sinai |
| Suzanne Sandmeyer | UCI/CHOC |
| Tahseen Mozaffar | UCI/CHOC |
| Tammi Skelton | UAB |
| Tanner D Jensen | Stanford |
| Tarun KK Mamidi | UAB |
| Taylor Beagle | Sanford |
| Taylor Maurer | Stanford |
| Teodoro Jerves Serrano | Yale |
| Terra R. Coakley | Stanford |
| Thomas Cassini | Vanderbilt |
| Thomas J. Nicholas | University of Utah |
| Timothy Schedl | WUSTL MOSC |
| Tiphanie P. Vogel | BCM Clinical |
| Vaidehi Jobanputra | Columbia |
| Valerie V. Maduro | NIH UDP |

| | |
|---|---|
| Vandana Shashi | Duke |
| Vasilis Vasiliou | Yale |
| Virginia Sybert | PNW |
| Vishnu Cuddapah | CHOP |
| Wendy Introne | NIH UDP, NHGRI |
| Wendy Raskind | PNW |
| Willa Thorson | Miami |
| William A. Gahl | NIH UDP, NHGRI |
| William E. Byrd | UAB DMCC |
| William J. Craigen | BCM Clinical |
| Winston Halstead | Yale |
| Yan Huang | NIH UDP, NHGRI |
| Yigit Karasozen | UCLA |
| Yong-Hui Jiang | Yale |
